# Redefining tissue specificity of genetic regulation of gene expression in the presence of allelic heterogeneity

**DOI:** 10.1101/2021.06.28.21259545

**Authors:** Marios Arvanitis, Karl Tayeb, Benjamin J. Strober, Alexis Battle

## Abstract

Understanding the mechanisms that underlie genetic regulation of gene expression is crucial to explaining the diversity that governs complex traits. Large scale expression quantitative trait locus (eQTL) studies have been instrumental in identifying genetic variants that influence the expression of target genes. However, a large fraction of disease-associated genetic variants have not been clearly explained by current eQTL data, frustrating attempts to use these data to comprehensively characterize disease loci. One notable observation from recent studies is that cis-eQTL effects are often shared across different cell types and tissues. This would suggest that common genetic variants impacting steady-state, adult gene expression are largely tolerated, shared across tissues, and less relevant to disease. However, allelic heterogeneity and complex patterns of linkage disequilibrium (LD) within each locus may skew the quantification of sharing of genetic effects between tissues, impede our ability to identify causal variants, and hinder the identification of regulatory effects for disease-associated genetic variants. Indeed, recent research suggests that multiple causal variants are often present in many eQTL and complex trait associated loci. Here, we re-analyze tissue-specificity of genetic effects in the presence of LD and allelic heterogeneity, proposing a novel method, CAFEH, that improves the identification of causal regulatory variants across tissues and their relationship to disease loci.

## introduction

Understanding the mechanisms that underlie genetic regulation of gene expression is crucial to explaining the diversity that governs complex traits. Large scale expression quantitative trait locus (eQTL) studies have been instrumental in identifying genetic variants that influence the expression of target genes. However, a large fraction of disease-associated genetic variants have not been clearly explained by current eQTL data [1,2], frustrating attempts to use these data to comprehensively characterize disease loci. One notable observation from recent studies is that cis-eQTL effects are often shared across different cell types and tissues [3-5]. This would suggest that common genetic variants impacting steady-state, adult gene expression are largely tolerated, shared across tissues, and less relevant to disease [6]. However, allelic heterogeneity and complex patterns of linkage disequilibrium (LD) within each locus may skew the quantification of sharing of genetic effects between tissues, impede our ability to identify causal variants, and hinder the identification of regulatory effects for disease-associated genetic variants. Indeed, recent research suggests that multiple causal variants are often present in many eQTL and complex trait associated loci [7]. Here, we re-analyze tissue-specificity of genetic effects in the presence of LD and allelic heterogeneity, proposing a novel method, CAFEH, that improves the identification of causal regulatory variants across tissues and their relationship to disease loci.

## Results

Previous analyses of cis-eQTL effect sharing across tissues have employed meta-analytic strategies that aggregate the association signals from multiple tissues [8-11]. A crucial pitfall of these analyses lies in handling LD. Specifically, if two distinct causal regulatory cis-eQTL variants acting in separate tissues are in even moderate LD with each other, those variants often falsely appear to be active in both tissues, boosting each-other’s association signal and providing an often false, high estimate of eQTL sharing between tissues (example, Figure 1A). Indeed, tissue sharing statistics reported by the GTEx Project [10], quantified by Metasoft [8] m-values, are strongly correlated with LD score (Figure 1B), indicating tissue sharing estimates are likely to be inflated by LD. This association remains after controlling for gene density and distance to the nearest transcription start site (Supplementary Figure 1).

**Figure 1.**
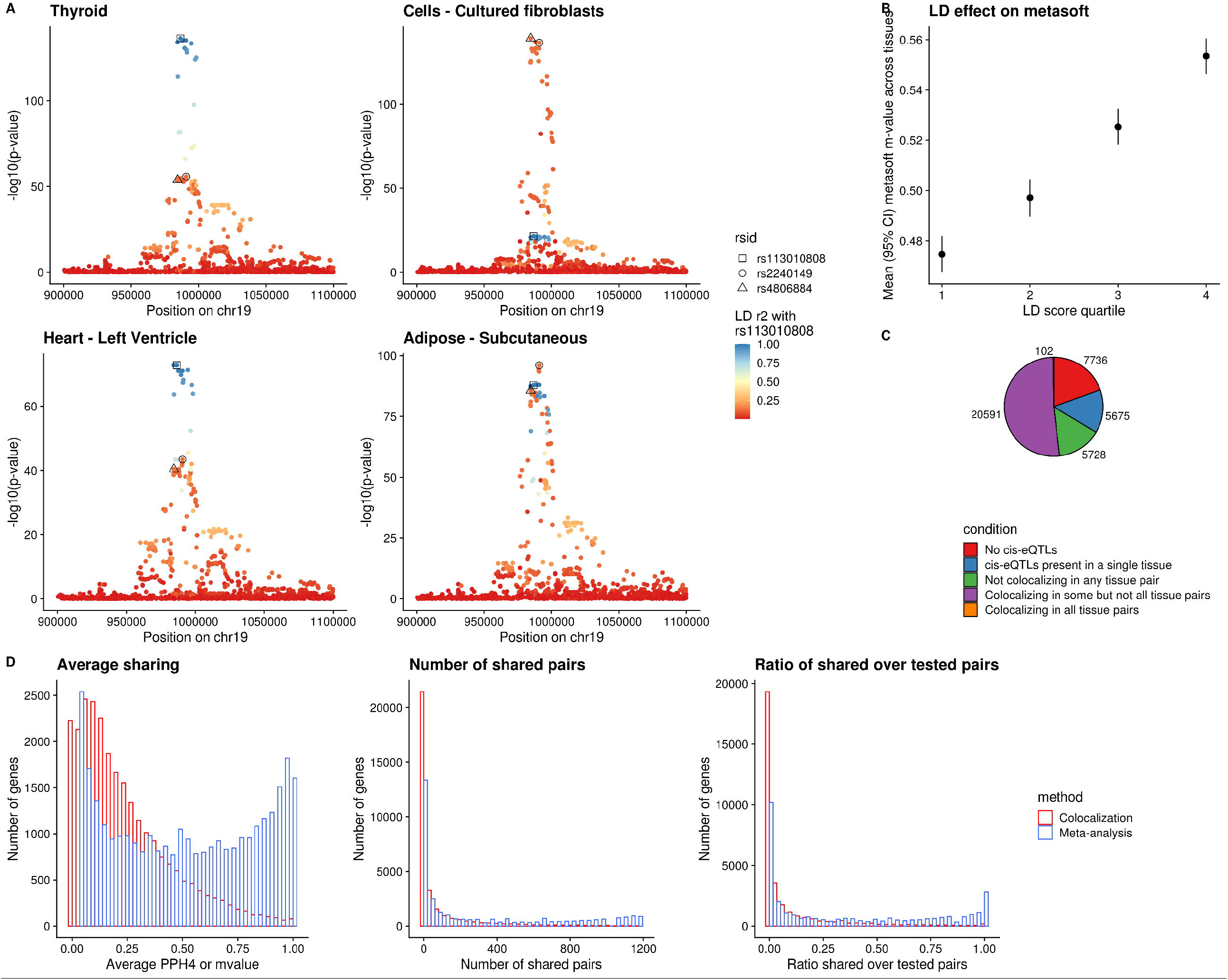
Colocalization provides evidence of extensive eQTL tissue specificity. **A**. Local Manhattan plots of cis-eQTLs for the WDR18 gene in four tissues of GTEx v8. The plots reveal allelic heterogeneity between tissues (while Thyroid and Heart share the same pattern of genetic regulation, Whole Blood and Fibroblasts have different causal variants). The three lead variants across the four tissues are colored based on their LD with variant rs113010808 (lead variant in both Thyroid and Heart). All three variants have a Metasoft m-value of 1 in all four tissues. LD r^2^ between rs2240149 and rs4806884 is 0.79 while r^2^ between rs2240149 and rs113010808 is 0.15. **B**. Metasoft m-values are positively correlated with LD. When the LD score quantile of the tested variant increases, the average m-value across tissues is higher. **C**. Pie-chart of all 38,518 genes in GTEx v8 that are expressed in at least one tissue, based on the tissue specificity of their eQTLs estimated by COLOC. **D**. Histograms depicting patterns of sharing of genetic regulation between tissues based on COLOC and Metasoft. From left to right: Histogram of mean PPH4 or m-value between tissue pairs for each gene; histogram of number of tissue pairs with shared cis-eQTLs for each gene; and histogram of the ratio of shared tissue pairs divided by the tissue pairs in which the gene is expressed. COLOC reveals more profound tissue specificity.

We performed an alternative analysis of tissue sharing among cis-eQTLs across 49 human cell types and tissues in GTEx v8 [10] using a colocalization approach (COLOC [13]) that explicitly incorporates LD information to overcome the confounding issues observed with other approaches and attempts to estimate shared causal variants. Colocalization has been commonly used for assessing the causal overlap between eQTLs and genome-wide association studies (GWAS), but infrequently used for assessing relationships between eQTLs [12]. Our analysis revealed that for the majority of genes, distinct causal variants are likely to be responsible for the cis-eQTL signals in different tissues (Figure 1C and 1D), with far less sharing than reported by meta-analysis approaches. Since COLOC is limited by the assumption of a single causal variant per tested region, we also performed another analysis using a second LD-aware colocalization method, eCAVIAR [14], allowing up to two causal variants per region. Because eCAVIAR is computationally expensive, it was run on a subset of 1000 randomly sampled genes. These results confirmed the pattern of widespread tissue-specificity in regulation of gene expression (Supplementary Figure 2A and 2B).

We found that gene-tissue pairs were identified as shared by Metasoft even when they had distinct top cis-eQTL variants in only moderate LD, whereas for colocalization, sharing was identified only when the lead variants were identical or in high LD with each other, more likely tagging the same causal effect (Figure 2A). Further, using colocalization, tissues with similar origin had a higher degree of sharing across genes, whereas with meta-analysis this expected pattern of sharing was far weaker, as evaluated across the full range of PPH4 or m-values, respectively (Figure 2B and Supplementary Figure 3). We should note that the overall degree of tissue sharing naturally changes depending on the selection of different thresholds for each method. For colocalization, even a remarkably lenient PPH4 threshold of 0.1 reveals substantial tissue specificity (Supplementary Figure 4). For Metasoft, higher m-value thresholds do shift to a more tissue-specific pattern (Supplementary Figure 5A), but primarily by selecting for eQTLs with a stronger p-value in each tissue (Supplementary Figure 6). Additionally, even at more stringent Metasoft thresholds, it does not identify the genes that appear to share effects through colocalization(Spearman ρ between COLOC PPH4 and Metasoft m-value = 0.57). Specifically, shared gene-tissue pairs identified by colocalization continue to show stronger LD between their lead variants compared to those identified as shared by Metasoft regardless of chosen threshold (Supplementary Figure 5B), suggesting more LD artifacts with Metasoft regardless of threshold. The observed cis-eQTL tissue specificity extends to datasets with larger sample size like the eQTLGen [6] and Muther [15] consortia (Figure 2C, Supplementary Figure 7). Estimates of sharing in bulk tissue samples with cellular heterogeneity are also affected by cell type composition variability; genes with a strong posterior probability for separate, independent signals between eQTLGen and GTEx whole blood tissue (PPH3>0.9) were enriched for cell-type interaction QTL signals compared to genes with colocalization between the two datasets (PPH4>0.9) (Figure 2D).

**Figure 2.**
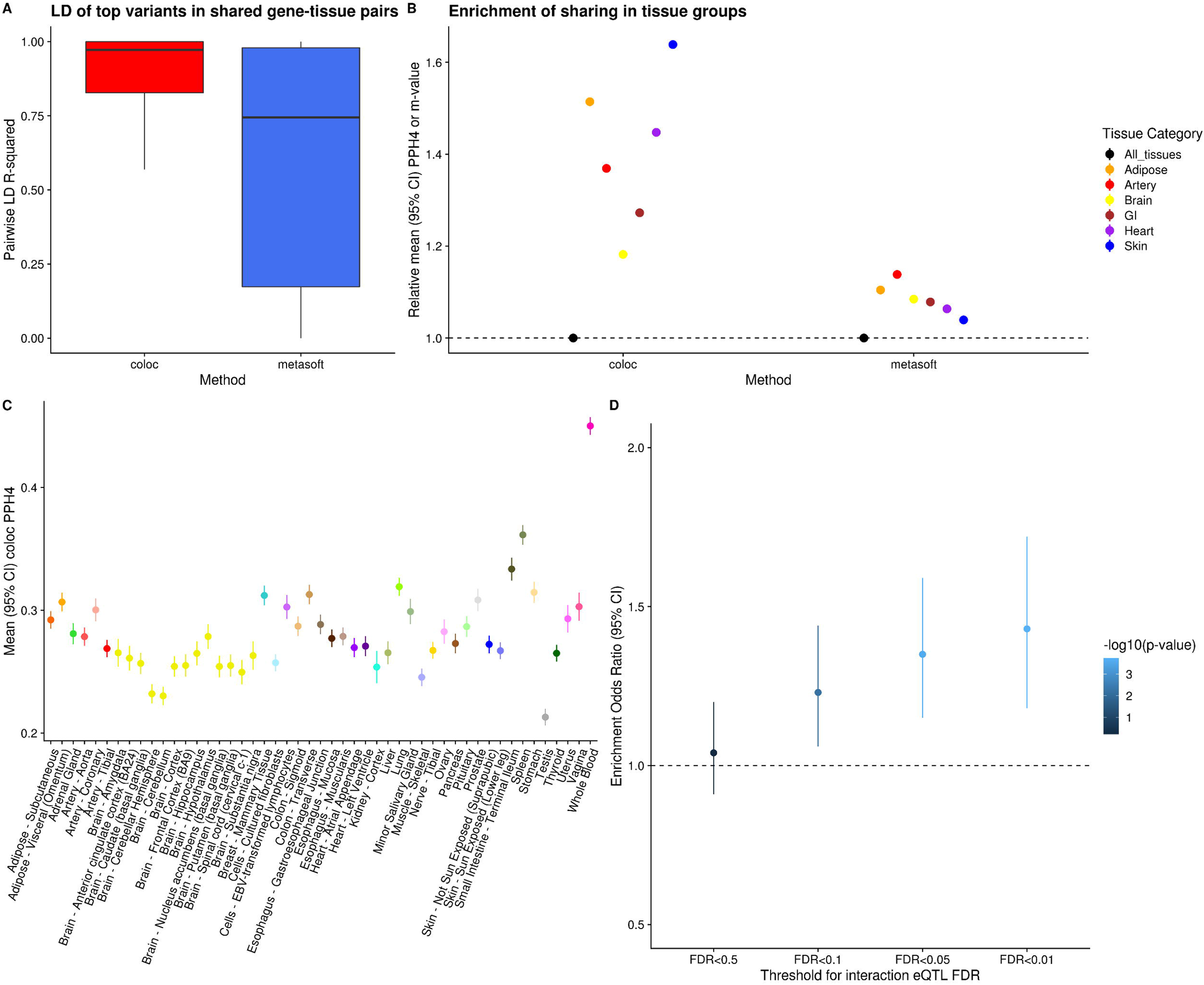
Colocalization is superior to other methods that don’t account for LD. **A**. Boxplots of the LD between the top variants per gene in tissue pairs that colocalize based on COLOC (in red) or Metasoft (in blue). **B**. Average sharing between groups of biologically similar tissues in GTEx v8 based on COLOC or Metasoft. Values are normalized by dividing with the mean across all gene-tissue pairs. **C**. Average eQTL sharing between eQTLGen Whole Blood and all GTEx v8 tissues for genes that have an eQTL in both datasets, using COLOC PPH4. **D**. Odds ratio of enrichment for non-colocalizing genes (PPH3>0.9) compared to colocalizing genes (PPH4>0.9) between eQTLGen Whole Blood and GTEx Whole Blood. The odds ratios are stratified by the false discovery rate of interaction with cell type composition, measured as the interaction between the lead eQTL variant for that gene in GTEx v8 and the relative proportion of neutrophils (the most common Whole Blood cell type) in the GTEx sample.

Existing methods for colocalization, despite accounting for LD better than meta-analysis approaches, have several known limitations which preclude full exploration of tissue-specificity and causal variant sharing. First, most existing colocalization methods require manual specification of the number of causal variants for each locus, and those that allow for more than one causal variant become computationally intractable when many causal variants are specified, thereby substantially limiting evaluation of allelic heterogeneity. Further, even when two studies have the same underlying LD, most colocalization methods underperform in regions of high LD in a manner biased against reporting colocalization [14]. Nonetheless, we should note that this bias is not enough to account for the observed cis-eQTL tissue specificity as the evidence of pervasive tissue specificity well beyond that reported by meta-analysis methods remains present across different underlying LD structures (Supplementary Figure 8). Last, most existing methods generally perform pairwise comparisons between studies, therefore failing to aggregate evidence from multiple studies or tissues jointly. All these limitations are highlighted in simulations we performed across a range of chosen number of causal variants, underlying LD structures, and different populations assessed (Supplementary Figure 9 and 10), which show suboptimal performance of colocalization in the presence of allelic heterogeneity and high LD by COLOC.

To overcome the limitations of existing approaches and to robustly perform fine-mapping and colocalization in the presence of multiple causal variants across many studies, we developed CAFEH (Colocalization And Fine-mapping under allElic Heterogeneity). CAFEH is a Bayesian algorithm that incorporates information regarding the strength of the association between a phenotype and the genotype in a locus along with LD structure of that locus across different studies and tissues to infer causal variants within each locus. Depending on availability of individual level genotype and phenotype data, we developed two versions of the method, CAFEH-G (Figure 3A), when individual level data is available, and CAFEH-S, that can be applied with summary statistics alone. CAFEH does not depend on pre-specified assumptions on the number of causal variants per locus, and outperforms existing methods both in precision-recall and computational efficiency as shown in simulations (Figure 3D and 3E and Supplementary Figures 11-14). In addition, CAFEH leverages the signal across multiple studies jointly, therefore increasing the accuracy of fine-mapping compared to existing, single-study approaches (Figure 3E).

**Figure 3.**
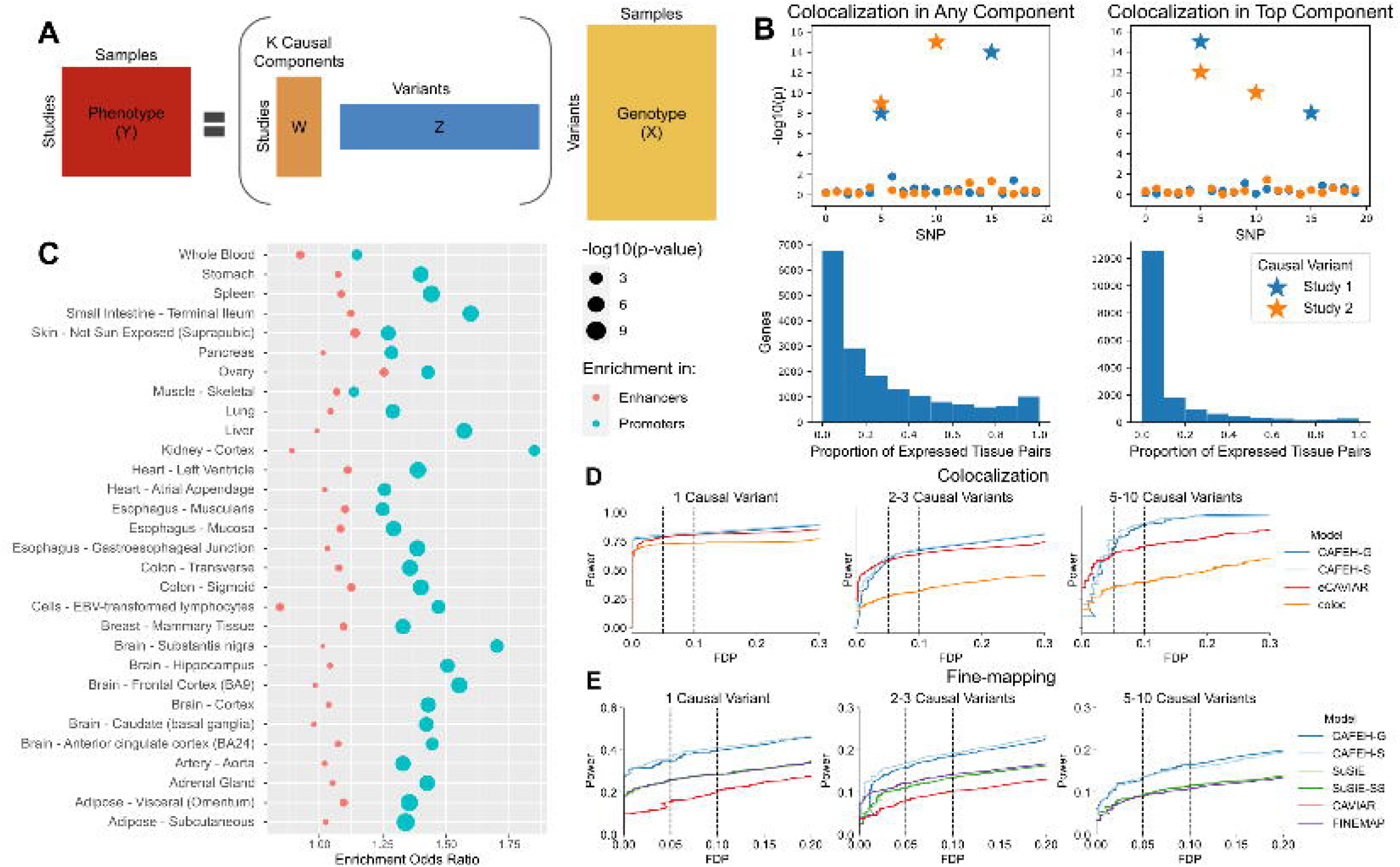
CAFEH GTEx and Simulations. **A**. A schematic representation of CAFEH. CAFEH can be viewed as a sparse regression with a shared set of causal variants across all studies. Entries of W are modeled with a spike and slab prior, so each study uses a subset of causal variants. **B**. Redefining colocalization with allelic heterogeneity. Top: representation of colocalization in any or the top component. Stars represent causal variants in each study. Bottom: Proportion of tissue pairs colocalizing in any or top CAFEH components at a 0.5 threshold. **C**. Enrichment of the top CAFEH variant of each gene in promoter (teal) and enhancer (red) elements in matched Roadmap cell-types relative to top eQTL variants. **D**. Proportion of false discoveries (FDP) vs proportion of true positives (Power) across a range of colocalization thresholds. **E**. FDP vs Power across a range of thresholds of the posterior inclusion probability in competing fine-mapping methods.

Importantly, when tested across 49 GTEx v8 tissues, CAFEH-G recapitulates the pervasive tissue specificity in genetic regulation of gene expression (Figure 3B and Supplementary Figure 15). Further, by identifying allelic heterogeneity within each locus, CAFEH allows us to redefine colocalization based on different patterns of sharing of causal variants. For example, two studies may share all causal variants from either study, a very stringent definition of colocalization. Alternatively, they may share their most strongly association causal component, or any subset of their active causal genetic components, a less stringent but still potentially informative form of colocalization (Figure 3B). Using the output from CAFEH, these, and other customized criteria may be applied to define colocalization by each user depending on the particular goals of their study (Online Methods). Notably, CAFEH-G reveals that the majority of genes have more than one causal variant per tested tissue (Supplementary Figure 16). Similarly, when tested across all 49 GTEx v8 tissues, most genes have five or more total different causal variants influencing their expression among all tissues (Supplementary Figure 15). Lastly, causal variants fine-mapped by CAFEH in each tissue are enriched for promoter elements in the corresponding tissues as defined by Roadmap chromHMM [16], compared to top eQTL variants for the corresponding genes (Figure 3C). We should note that by nature of its joint cross-tissue inference, CAFEH prefers components that have evidence for an active signal in multiple studies and hence has a slight bias towards colocalization. However, that is offset by the increase in power that the joint inference allows, hence improving overall accuracy in both fine-mapping and colocalization (Figure 3D and 3E and Supplementary Figure 13). We should also note that despite this potential bias towards sharing, CAFEH still reveals substantial evidence for tissue specificity of cis-eQTLs.

We observed that genes whose genetic transcriptional regulation is predominantly tissue specific as defined by CAFEH have different characteristics than those whose regulation is shared across tissues. First, patterns of evolutionary conservation were different between the set of genes that are highly shared between tissues and those that are predominantly tissue specific, restricting to genes that have a significant cis-eQTL in at least 20 tissues each. Specifically, tissue specific genes were found to be more variation intolerant as measured by pLI [17] compared to background genes, which were in turn more intolerant compared to the subset of genes with shared genetic regulation (Figure 4B), a phenomenon present regardless of the underlying strength of the eQTL association (Supplementary Figure 17). Further, gene ontology (GO) enrichment analysis [18] demonstrated that poorly colocalizing genes were enriched in pathways related to development, differentiation, cell-adhesion and transcription factor activity, suggesting that these pathways are critical to the differences between cell types in humans despite being broadly expressed and genetically regulated in many tissues. In contrast, highly colocalizing genes were enriched for mitochondria and the cytosolic ribosome, cellular structures that are abundant in all tissues and cell types (Figure 4A). In parallel, we observed that poorly colocalizing genes were enriched for participation in human diseases with dominant inheritance patterns as defined by OMIM [19] (Figure 4C). We then showed that causal eQTL components shared by very few tissues are more likely to be active in GWAS than those shared by multiple tissues (Figure 5A). Conversely, GWAS causal components that are also eQTLs in known target tissues are more likely to be tissue specific compared to general eQTL causal components in those tissues (Figure 5B). These results are consistent with our finding of richer evolutionary conservation in tissue specific compared to tissue shared genes and jointly suggest that exploration of cell type specific and potentially context specific eQTLs could provide an explanation of the effects of GWAS variants that lack a colocalizing signal in currently available eQTL datasets.

**Figure 4.**
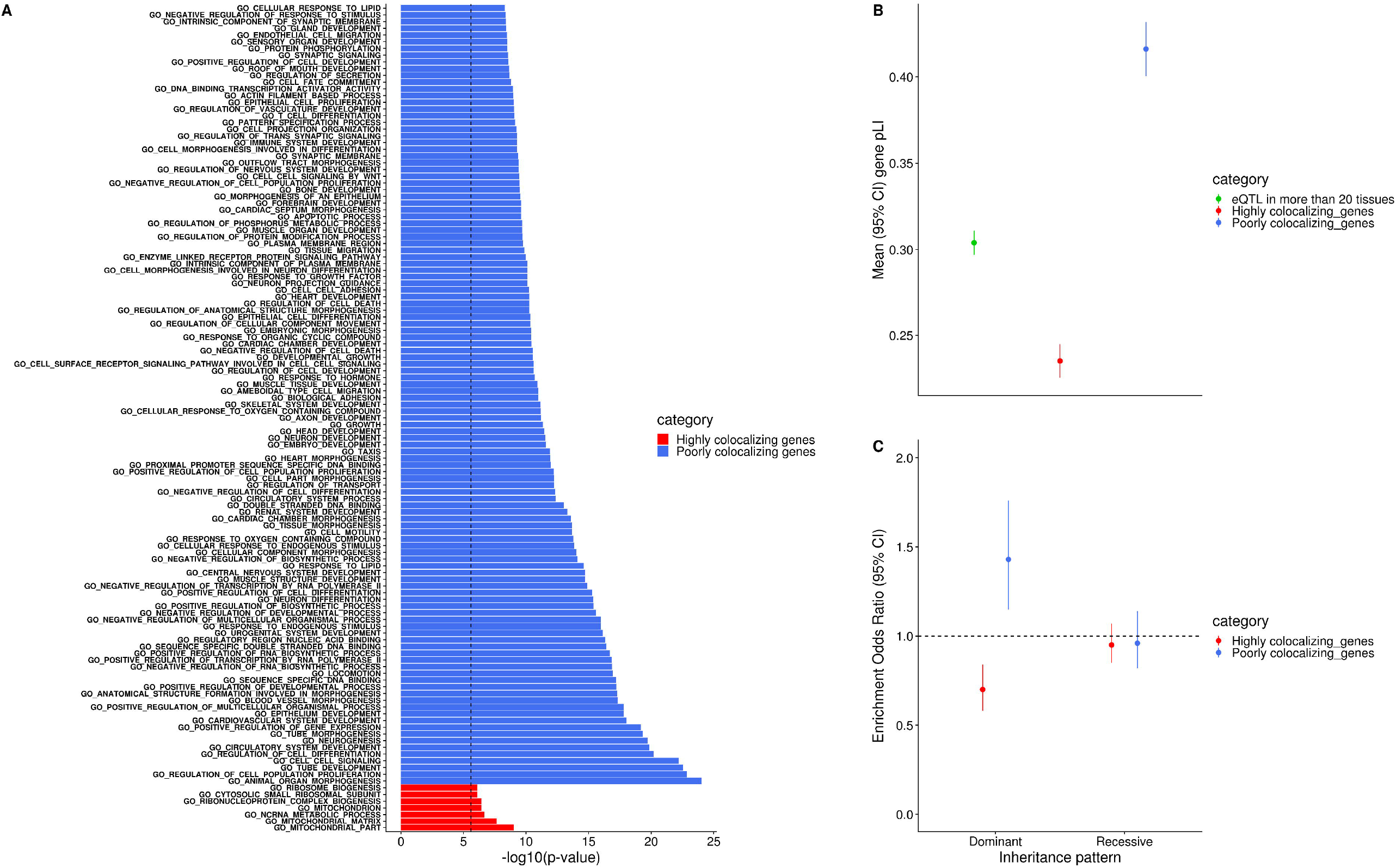
Characteristics of genes based on the degree of eQTL tissue sharing. **A**. Gene Ontology enrichment analysis of genes that are colocalizing in at least 20 tissues (highly colocalizing) and those that colocalize in less than 5 tissues (poorly colocalizing), comparing only genes that have an eQTL in at least 20 tissues. Colocalization was defined as sharing of the top causal component based on CAFEH. **B**. Average probability of loss of function intolerance (pLI, left panel) between the same three groups of genes as panel A. **C**. Enrichment in genes whose rare variation is associated with autosomal dominant or recessive human diseases based on OMIM (left panel) between highly and poorly colocalizing genes.

**Figure 5.**
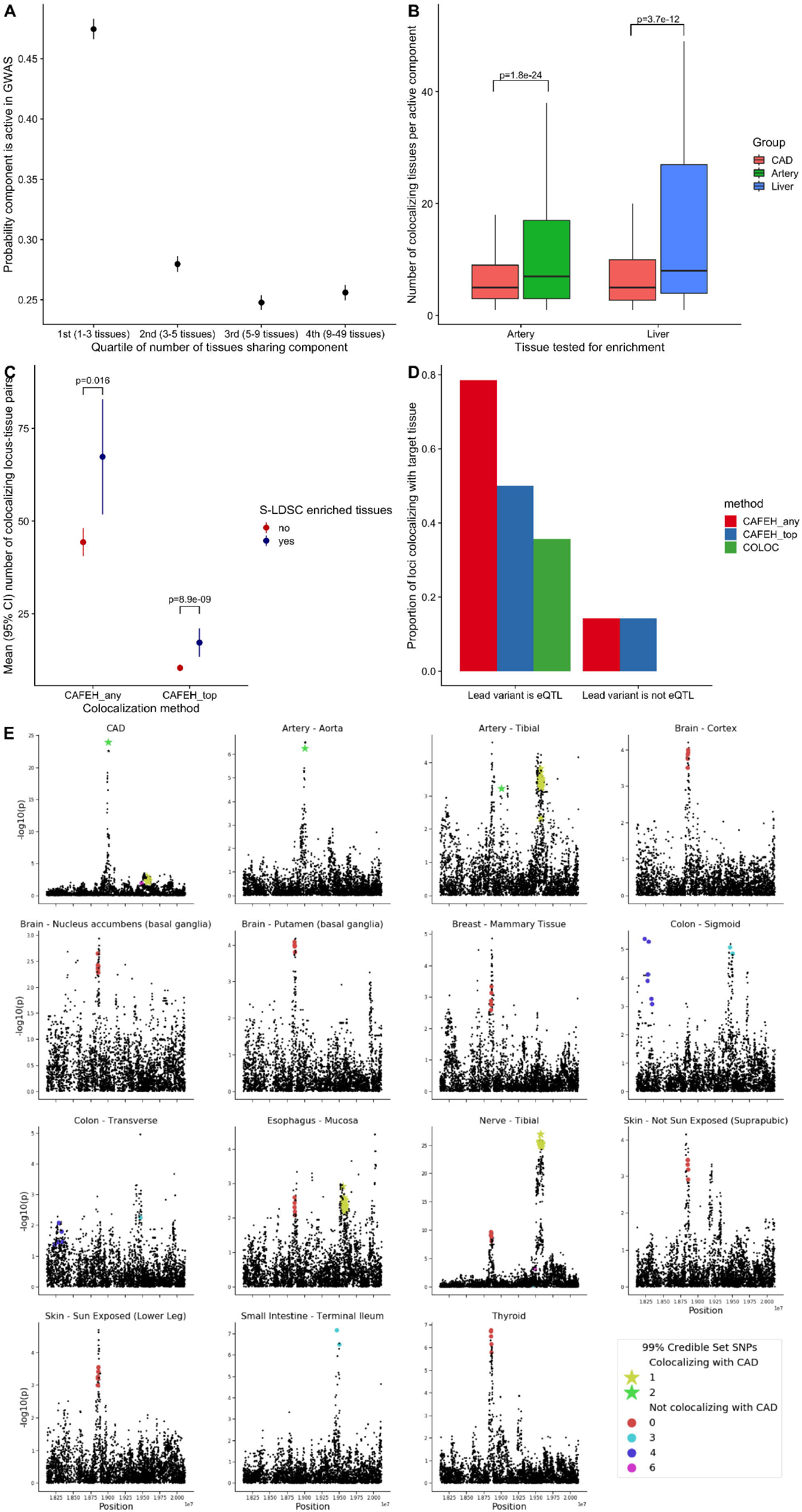
CAFEH identifies the target tissue in GWAS. **A**. Mean (95% CI) of probability of a cis-eQTL component being active in GWAS (as defined by CAFEH-S) stratified by the quartile of the number of 49 GTEx tissues that share the given eQTL. The figure presents aggregate results of 19 diverse GWAS traits from the UK Biobank. For each GWAS, we jointly run CAFEH-S between the GWAS summary statistics and GTEx v8 eQTL summary statistics among genes for which the sentinel variant of a genome-wide significant disease locus is also a genome-wide eQTL in at least one tissue. We then evaluate all components that are active in at least one GTEx tissue. Overall there is a strong inverse association between the posterior probability of a component being active in the GWAS and the number of tissues that colocalize with said component (linear mixed model p = 3.3e-160). **B**. Boxplots of the number of tissues that share an active component identified by CAFEH, among components that are either active in a CAD GWAS and a GTEx tissue a priori believed to be relevant for CAD heritability (either Artery on the left or Liver on the right) among all genes within 1Mb of genome-wide significant CAD locus, compared to components that are active only in a GTEx tissue (Artery or Liver) among all protein coding genes. **C**. Number of colocalizing locus tissue pairs in 19 diverse GWAS traits from the UK Biobank. Colocalization is defined based on any component or top component in CAFEH and the counts are colored based on whether the corresponding tissues are enriched for that trait by partitioned LD score regression. **D**. Case study of loci in a large CAD GWAS that have an established target tissue. The bars represent the proportion of loci colocalizing in the known target tissue with any of three different methods (CAFEH colocalization in any component, CAFEH colocalization in the top component, COLOC). The results are stratified based on whether or not the lead variant in the locus is a genome-wide significant cis-eQTL for any gene in at least one tissue in GTEx v8. We see that CAFEH outperforms COLOC and can prioritize the target tissue in most cases when an eQTL signal is present. **E**. Example of CAFEH revealing allelic heterogeneity and identifying the target tissue in the TWIST1 CAD locus. CAFEH identifies a dominant component for the CAD GWAS in that locus with a 99% credible set that consists of a single variant (rs2107595). The same component is shared by artery tissue in GTEx v8. Recent CRISPR in human arterial smooth muscle cells confirmed its effects in influencing the expression of TWIST1 in that tissue.

The underappreciated yet extensive tissue specificity in cis-eQTLs should also in theory provide a basis to probe the tissue of interest in genome-wide association study (GWAS) loci for a number of complex traits. Although researchers have increasingly relied on colocalization to identify gene candidates for significant GWAS associations [20], previous attempts to define the target tissue based on eQTL data have shown some promise [21] but investigators have not recommended the use of eQTL information in prioritizing target tissues in GWAS [21,22] citing the tissue-sharing of cis-eQTLs in a large fraction of trait associations as one reason [23].

However, the predominant tissue specificity that we have observed in CAFEH causal components suggests that when a colocalizing eQTL signal is present within the GWAS locus, CAFEH can leverage its tissue specific regulation to improve accuracy in identifying target tissues in which said component exerts its effect. Indeed, we showed that CAFEH-S accurately prioritizes the target tissue for a number of diverse traits from the UK Biobank and the tissue prioritization results agree with stratified LD score regression estimates applied to the same traits (Figure 5C, Supplementary Figure 18). Next, we evaluated whether CAFEH-S is able to prioritize the correct tissue in GWAS variants where the active tissue is known. To do that, we performed a case study in coronary artery disease (CAD), a sample complex trait with substantial heritability [24] and a pathophysiology that involves a variety of different organ systems [25]. We assessed the performance of CAFEH-S in identifying the causal tissue for all CAD loci that have either been subjected to experimental validation of the causal tissue via genome editing or loci in which the closest gene is an established CAD gene with a known tissue-specific mechanism of action. CAFEH-S colocalizes with the correct tissue in those loci 80% of the time and outperforms COLOC when a significant eQTL signal is present, demonstrating that the approach is effective in most situations (Figure 5D and 5E, Supplementary Figure 19) when highly powered GWAS and corresponding tissue eQTL summary statistics are available. Naturally, although the method identifies colocalization even in weak (non-genome-wide significant) eQTL signals (see SMAD3 locus in Supplementary Figure 19), it is unable to colocalize with the target tissue when the eQTL signal is absent. We should note that of 6 loci without a genome-wide significant eQTL signal, three (ANGPTL4, APOE, LDLR) contain a coding variant of the corresponding target gene in high LD (R2 > 0.6) with the sentinel SNP which may suggest their effects on phenotype may not be mediated by gene expression. Lastly, as we have shown here and previous studies support [26, 27], a broader set of eQTL data from specific cell types and different contexts is likely to improve our ability to identify the cell type of interest across different GWAS traits.

In summary, our results provide evidence of pervasive tissue specificity in genetic regulation of gene expression. We develop a new computational tool, CAFEH, to perform fine-mapping and colocalization jointly across multiple tissues and traits that allows multiple causal variants present across studies and can better explore sharing in the presence of allelic heterogeneity. We use that tool to show that genes whose transcriptional regulation is tissue specific, despite being broadly expressed and genetically regulated, tend to be conserved and more relevant in disease and that causal GWAS signals are more likely to be tissue specific than shared. Finally, we demonstrate that the method is effective at leveraging the tissue specificity of eQTLs to improve the identification of target tissue in GWAS loci.

## Online Methods

### Evaluation of tissue sharing in GTEx v8

The primary source of data for this analysis were eQTL summary statistics across 49 human tissues and cell types generated by the GTEx Consortium v8 release [10]. We additionally used individual level whole genome and RNA-sequencing by GTEx processed according to the GTEx v8 protocol [10]. Metasoft analysis was performed as previously described in GTEx v8 [28]. To perform colocalization we employed COLOC with the approximate Bayes method [13] for each gene locus defined as all SNPs in a region within 1 Mb from the corresponding gene transcription start site. COLOC was performed in a pairwise manner between all 49 GTEx v8 tissues for all genes that were expressed in at least one tissue in GTEx v8. Priors for the different colocalization probabilities were set at p1 = 1* 10^−4^, p2 = 1* 10^−4^, p12 = 1* 10^−6^according to the authors’ recommendation in the original COLOC paper [13]. Colocalization was defined as a PPH4 ≥ 0.5 unless explicitly stated otherwise. Gene-tissue pairs that did not have a signal for an association with genotype in both tested studies (ie. gene-tissue pairs with PPH3+PPH4 < 0.5) were excluded from the analysis of tissue sharing.

Since COLOC evaluates the probability of colocalization at the locus level considering the locus signal as a whole, whereas Metasoft m-values work on the variant level and determine the probability for a given variant to be significant eQTL for a gene-tissue pair, in order to perform comparisons between the two we defined that two tissues shared regulation for a given gene based on Metasoft if the m-value for the variant with the minimum eQTL p-value across all GTEx tissues for that gene was ≥ 0.5 in both tissues.

COLOC by default assumes at most a single causal variant per locus. That assumption may influence the results in loci with multiple causal variants biasing them against colocalization. Therefore, to test whether the substantial tissue specificity observed by COLOC was significantly influenced by that limitation, we repeated our colocalization analysis using eCAVIAR [11], a software that relaxes the single causal variant assumption. A limitation of eCAVIAR is that it scales exponentially in the number of assumed causal variants making inference of colocalization substantially slower as the number of variants increases. Consequently, even assuming at most two causal variants per-locus, this analysis was not computationally feasible across all 35,848 genes and 49 tissues evaluated with COLOC. Therefore, we performed eCAVIAR assuming at most two causal variants per locus in a randomly sampled subset of 1,000 genes out of the 35,848 to evaluate the distribution of genes with shared and distinct regulation in different tissues pairwise. We defined that two tissues colocalize based on eCAVIAR if they share at least one variant with a minimum causal posterior probability in both tissues ≥ 0.5.

### Evaluation of tissue sharing between GTEx and other datasets

We subsequently evaluated tissue sharing between all 49 tissues in GTEx v8 and human tissues or cell types from Muther [15] and eQTLGen [6]. Specifically, for each of the three Muther tissues and cell types that cis-eQTL data are available (Fat, Skin and lymphoblastoid cell lines), we performed pairwise colocalization analysis for all genes that were tested in both Muther and GTEx v8 between the corresponding Muther tissue and all 49 GTEx tissues. We plotted the colocalization posterior probability (PPH4) mean and 95% confidence intervals using the subset of gene-tissue pairs that had an eQTL signal in both tissues based on the COLOC output (ie. PPH3+PPH4 ≥ 0.5). We followed the same procedure to test for colocalization between eQTLGen whole blood and all 49 GTEx tissues.

### Cell type deconvolution in GTEx Whole Blood

We performed cell type deconvolution analysis in GTEx whole blood tissue using CIBERSORT [29]. Specifically, we ran CIBERSORT with default parameters using as input the GTEx v8 whole blood expression in transcripts per million and the signature genes and average expression from a dataset of 22 circulating human immune cell types [29]. We classified a cell type as estimable in GTEx if it had a corresponding CIBERSORT estimate > 0.05% in more than 5% of the GTEx whole blood samples [30]. We were able to estimate cell type proportions in GTEx for 15 different immune cell types using the above method.

After obtaining the CIBERSORT estimates, we performed interaction QTL calling using MatrixEQTL [30] and the following linear model:

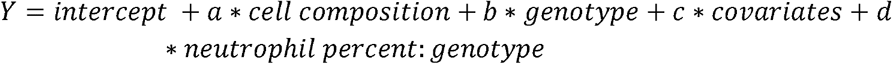

Where *Y* is the processed gene expression, *genotype* is the genotype of the lead cis-eQTL SNP for that gene, *cell composition* is a matrix containing the cell type proportions for each of the 15 estimable cell types in GTEx, covariates include sex, PCR, platform, 60 PEER factors and 5 genotype PCs and *neutrophil percent: genotype* is the interaction term between the SNP genotype and the proportion of neutrophils in each GTEx sample. We evaluated statistical significance of the interaction effect estimate *d* using a two-sided Wald test and performed Benjamini-Hochberg correction of the p-values across tested genes. We then selected the genes that had a significant interaction QTL at an FDR < 0.05 and tested whether genes that had high posterior probability for non-colocalization between eQTLGen and GTEx whole blood (PPH3 > 0.9) would be enriched in genes with a significant interaction QTL compared to genes that had high PPH4 (suggesting that cell type differences influence colocalization estimates between the two datasets).

### Simulations to evaluate COLOC performance

To evaluate the performance of COLOC under different underlying LD patterns and numbers of causal variants in each locus, we performed a series of simulations. To ensure we have a broad representation of LD structures in our simulations, we first computed LD scores for all variants in GTEx v8 whole genome sequencing that passed the standard GTEx v8 filters using the ldsc software [31]. Naturally, genes with higher LD score are expected to be found in regions with higher LD on average. We then split each gene that had a significant cis-eQTL in GTEx whole blood into LD quintiles based on the LD score of its top eVariant. Subsequently, we randomly sampled 20 different genes from each LD bin. For each gene we obtained genotypes for the corresponding locus by selecting the SNPs within 1Mb from the gene’s transcription start site in four different datasets:

1. GTEx whole blood
2. GTEx thyroid
3. 1000 Genomes Europeans and
4. 1000 Genomes Africans.

We evaluated three possibilities regarding the number of causal variants:

1. There is a single causal variant which was selected to be the top cis-eVariant for the corresponding gene in GTEx whole blood
2. There are two causal variants, one of which is the top cis-eVariant for the corresponding gene in GTEx whole blood and the others are selected randomly among the remaining locus variants
3. There are five causal variants, one of which is the top cis-eVariant for the corresponding gene in GTEx whole blood and the others are selected randomly among the remaining locus variants

For all configurations, we simulated gene expression using the following linear model:

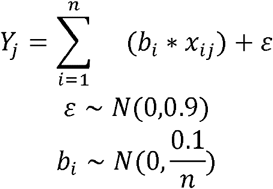

Where *Y*_*i*_ is the simulated expression for the *j*_*th*_ individual, *n* is the number of causal variants, *b*_*i*_ is the effect size for the *i*_*th*_ causal variant, assumed to have a normal distribution with heritability 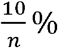, according to our prior knowledge on average cis heritability of gene expression [14], *x*_*ij*_ is the genotype for causal variant *i* and individual *j*, and *ε* is the residual error term with a normal, zero-mean distribution.

For each run of the simulation, we simulated gene expression using the above method for GTEx whole blood individuals and one of the four different genotype datasets listed above. After simulating gene expression, we then obtained simulated eQTL summary statistics for each variant in a locus by performing simple linear regression between the simulated gene expression and the variant genotypes. COLOC was performed on the simulated summary statistics between GTEx whole blood and each dataset using the same priors as outlined above for the analysis of real data. 100 independent simulations were performed for each dataset and causal variant configuration.

### CAFEH

CAFEH is a probabilistic model that performs colocalization and fine mapping jointly across multiple phenotypes. Let *Y* be an *N × T* matrix of measurements from *N* individuals in *T* phenotypes. Let *X* be an *N × G* matrix of genotypes for each individual in *G* SNPs. We assume an additive genetic model

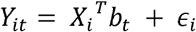

Where *b*_*t*_ is a sparse vector of effect sizes in phenotype *t*, and *∈* _*i*_ ∼ *N*(0,*τ* ^-1^ is i.i.d. noise. We model *b*_*t*_ as

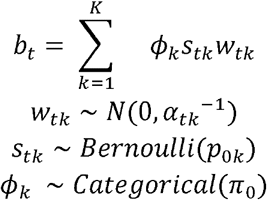

The model induces sparsity in two distinct ways that correspond with fine-mapping and colocalization respectively. First, *b*_*t*_ has at most *K* non-zero entries selected by the *K* categorical variables *ϕ*_*k*_. Similar to SuSiE [32], *b*_*t*_ is written as a sum of components where each component captures the effect of a single causal variant. Variants that don’t have a causal effect on any phenotype have by definition *b*_*t*_= 0. Therefore, inference about which variants have non-zero effect constitutes fine-mapping.

The *ϕ*_*k*_ are shared across all *T* phenotypes, that is, CAFEH models all *T* phenotypes with a common set of causal variants. Since we do not expect all causal variants to be relevant to all phenotypes, the effect sizes of each component in each phenotype are also sparse, modeled with a spike and slab prior parameterized by the product *s*_*tk tk*_. In particular, the *s*_*tk*_ are binary variables that select a subset of the *K* components to be active in phenotype *t*. Two phenotypes *t*_1_ and *t*_2_colocalize in component *k* if they are both active in component *k*, that is 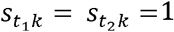.

To complete our model specification, we place priors on the variance terms for our effect sizes and residuals.

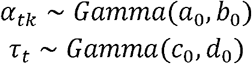

When *T*=1 CAFEH reduces to SuSiE with a spike and slab prior on the effect sizes. CAFEH generalizes SuSiE by sharing causal variants across multiple phenotypes. This enables straightforward colocalization analysis, and dramatically improves power to perform fine mapping of shared causal variants by sharing information across multiple phenotypes.

We refer to this form of the CAFEH model as CAFEH-G, to emphasize that it is fit with individual level genotype data.

### Fitting CAFEH from summary statistics

To facilitate the application of CAFEH to GWAS with publicly available summary statistics we implement a version of CAFEH, CAFEH-S, that can be estimated with summary statistics and a reference LD matrix, using the RSS likelihood [32]. The RSS likelihood relates the coefficients of a multivariate regression to the effect sizes and standard errors of the marginal univariate regressions. Let 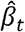 and 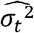be vectors of effect sizes and standard errors from a simple linear regression of phenotype *t* against a set of *G* SNPs. Let 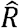 be the sample LD matrix computed on *X*. Define 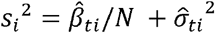 and *Ŝ* a diagonal matrix with *i*th diagonal equal to *s*_*i*_. Up to a constant, the likelihood *p* (*Y*_*t*_| *x, b*_*t*_)is equal to the likelihood of 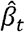 under the model (Proposition 2.1 in [39]):

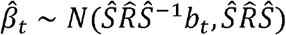

Thus we can equivalently do inference with summary statistics. In practice the sample LD matrix may not be available, and 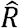 will need to be estimated from a panel of reference genotypes.

### Variational Inference for CAFEH

The exact posterior distribution *p* ({*w* _*tk*_},{*s*_*tk*_}, {*ϕ*_*k*_}, {*α* _*tk*_}, {*τ*_*t*_} | *Y, X*) is intractable, so we approximate the posterior distribution using variational inference. We select a family of distributions *Q* over the latent variables of the model that factorize as

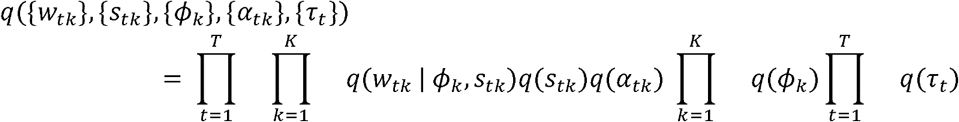

We perform coordinate ascent variational inference [33] to find a member of this variational family that (locally) minimizes the KL divergence to the true posterior distribution. All updates can be written in closed form. Detailed derivation of the updates for CAFEH-G and CAFEH-S, as well as implementation and initialization details are available in the Supplementary Methods, Section 2.4 and 2.3, respectively.

For CAFEH-S, to avoid costly matrix-vector multiplications at every iteration, we implement stochastic variational inference using a Monte-Carlo estimate of the variational objective. Specifically, we approximate expectations over *q* (*ϕ*) by sampling from Details are available in Supplementary Methods, Section 3.3.

### Setting hyperparameters

CAFEH users need to specify the number of components *K* and the *p*_*0k*_the prior probability that each component is active in each phenotype. *K* can be set to a large value (e.g. 20, 100) which is an upper bound on the number of causal variants CAFEH can detect. Irrelevant components will not be assigned to phenotypes. Similar to SuSiE, unused components have their posterior mass spread over a large number of variants so they do not significantly impact the posterior inclusion probabilities.

### Simulations

We compare the performance of CAFEH-S and CAFEH-G to popular fine-mapping methods including CAVIAR, FINEMAP, and SuSiE, and competing colocalization methods eCAVIAR and coloc. In all simulations CAVIAR and eCAVIAR are fit with a maximum of two causal variants, SuSiE with a maximum of 10 causal variants, and FINEMAP with a maximum of 5 causal variants. To better understand the impact of the spike and slab prior on fine-mapping we run CAFEH in each simulated phenotype separately, which we denote as (SuSiE-SS). We evaluate fine-mapping methods using the posterior inclusion probabilities (PIPs) returned by each model. We evaluate colocalization using colocalization statistics of each method, PPH4, CLPP, and p_coloc for coloc, eCAVIAR, and CAFEH respectively.

Gene expression data is simulated from real genotypes from 838 individuals in GTEx. We select 100 genes at random and take *X*^(*i*)^ to be the genotype matrix for the *G* variants nearest the transcription start site (TSS) of gene *i*.

Simulated expression is controlled by four parameters: *k* the number of causal variants in each phenotype, *ρ*the percent variance explained by causal variants, *r*^2^_*max*_ the maximum pairwise *r*^2^ between causal variants, and *n* the number of phenotypes simulated. Effect sizes are drawn from *N*(0,1/*p* (1 - *p*)where *p* is the allele frequency of the causal variant, residual variance is added to achieve the prescribed percent variance explained *ρ*. In each simulation we simulate multiple studies randomly assigned to two groups that share all their causal variants, and have distinct causal variants from the phenotypes in the other group.

In the main set of simulations, we simulate *n*= 4 studies, *r*^2^_*max*_ - 0.8, taking all combinations of *k*= 1,2,3 and *ρ* = 0.01,0.05,0.1,0.2 with *G* = 1000. We also generate expression for *k*= 5,10 and *ρ* = 0.2 using all variants within 1Mb of the TSS. The results of these simulations are highlighted in Figure 4D and 4E. eCAVIAR becomes computationally intractable on the larger simulation over the full cis-region, so for that simulation scenario, we run eCAVIAR using only variants with z score > 2 in at least one study. Across a range of signal strengths and genetic architectures CAFEH has more power to detect colocalizing phenotypes. While CAFEH’s performance is comparable to eCAVIAR when there are few causal variants, CAFEH performs much better when there is extensive allelic heterogeneity. We also note that CAFEH is able to vastly improve fine-mapping by performing fine-mapping jointly across all studies, compared to other methods that fine-map each phenotype separately. Compared to SuSiE, CAFEH’s 95% credible sets are smaller and detect a higher proportion of causal variants (Supplementary Figure 12).

To further investigate the value of fine-mapping shared causal variants jointly, we generate simulations where causal variants are shared across an increasing number of studies. In particular, we generate simulations with *n* = 4, 8,16 randomly assigned to two groups with *k*= 1,2,3 causal variants and *ρ* = 0.05. This simulation provides a range of scenarios where causal variants are shared between 1-12 phenotypes. CAFEHs PIPs better prioritize shared causal variants (Supplementary Figure 13)

While CAFEH is better able to fine-map variants that are shared across multiple phenotypes, it is also possible for CAFEH to represent multiple tightly linked causal variants with a single component, leading to false positive colocalization. To highlight this potential limitation, we generate simulations with *n*= 4, *k*=1 and *ρ* = 0.1. We vary the *r*^2^ between the causal variant in each group of studies in the ranges (0,0.5), (0.5,0.7), (0.7,0.9). We observe that CAFEH fails to distinguish between causal variants with *r*^2^ > 0.7

### Redefining colocalization with CAFEH

By design, CAFEH outputs credible sets of variants in each component identified as active in at least one tested study for a locus. Since each locus can (and often does) have more than one active component, there are many ways in which to define colocalization between two studies. For our analyses we chose the following three approaches (although other combinations can also be entertained):

1. Colocalization in any component: Defined as two studies sharing at least one component which is active in both studies with probability >=0.5.

2. Colocalization in the top component: Defined as two studies sharing their top component. To select a top component for each study, we generated a weight for all variants in the 95% credible set of all active components in the study defined as follows:

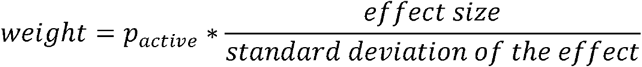

Where *p*_*active*_ is the probability of the component being active in the study, *effect size* is the effect size of the variant in the component and *standard deviation of the effect* is the standard deviation of that effect. We subsequently labeled as top component for each study the component that contains the variant with the maximum absolute value of the weight across all variants in the 95% credible sets of all components.

### Enrichment of CAFEH components in active regulatory elements

To evaluate the ability of CAFEH to identify causal variants in real data, we performed an enrichment analysis for variants in regulatory elements. Specifically, for each protein coding gene-tissue pair in GTEx, we selected the credible set variant that has the maximum weight as defined above (see top component colocalization) for that tissue. For each tissue we then compared the variants selected by this approach for each gene and evaluated for enrichment of those variants compared to a background of the variants that had the minimum cis-eQTL p-value for association with the expression of each gene in that tissue. The approach ensures that the same number of variants are included in the test and background sets since one variant is selected for each gene-tissue pair. We then evaluated for overlap between these variant sets and active regulatory elements in corresponding tissues from Roadmap Epigenomics [34] as defined in the main GTEx v8 paper [10] and performed a Fisher’s exact test to evaluate enrichment of the test compared to the background variant set.

### Gene set enrichment analysis

To evaluate whether genes with highly shared or very tissue specific regulation evaluated by CAFEH have distinct characteristics we performed gene set enrichment analysis based on sets defined by Gene Ontology (GO) [35] obtained by MSigDb [36]. Our background set of genes consisted of all protein-coding genes in GTEx v8 that have a significant cis-eQTL in at least 20 tissues. We tested two sets of genes against the background: a highly colocalizing set, defined as the subset of background genes that are colocalizing in their top component in at least 20 tissues, and a poorly colocalizing set, defined as the subset of background genes that are colocalizing in their top component in < 10 tissues. Fisher’s exact test was used for the gene set enrichment analysis for each GO term and a Bonferroni correction was applied for multiple testing. GO terms that had a Bonferroni-adjusted p-value < 0.05 were defined as enriched.

Similarly, the same sets of genes were evaluated for enrichment in sets of genes identified by OMIM as associated with human Mendelian diseases. We separated OMIM genes in two groups based on the underlying patterns of inheritance: Genes with autosomal dominant inheritance and genes with autosomal recessive inheritance.

We also evaluated the relative evolutionary conservation status of the same set of genes using pLI as a measure of conservation (higher pLI means that the gene is more conserved). We compared the pLI distributions between the two gene sets and the background set using a Wilcoxon rank sum test.

### Using CAFEH to infer sharing and target tissue in GWAS loci

We used CAFEH to evaluate the degree of tissue sharing in causal GWAS signals. We first (Figure 6A) evaluated 19 highly powered GWA studies from the UK Biobank [37]. All GWA loci were evaluated for each study. For each individual run of CAFEH, the input was one of the GWAS loci and the cis-eQTL summary statistics of all genes that are significant cis-eGenes for the sentinel variant in the GWAS locus across all 49 GTEx v8 tissues. GTEx v8 LD was used as reference. For each CAFEH run, we extracted all components identified as active by CAFEH in at least one GTEx v8 tissue (at a threshold of 0.5). We then stratified those components in quartiles based on the number of GTEx v8 tissues that share the component and we evaluated the mean CAFEH posterior probability of the component being active (p_active) in the GWAS in each quartile. We also evaluated for the overall association between the p_active in GWAS and the number of tissues that share said component by performing a linear mixed model with the GWAS phenotype as the random effects term and the number of tissues sharing the component as the fixed effect. The model was the following: p_active ∼ number_tissues_sharing_component + (1|phenotype).

We then evaluated whether components active in GWAS are more likely to be tissue specific than general eQTLs (Figure 6B). For that, we used a published GWAS meta-analysis of CAD [24] which is a disease with high heritability that is known to be enriched for the liver and the arterial wall. We then ran CAFEH-S jointly for all genes within 1Mb of each sentinel GWAS variant, between the GWAS and all tissues in GTEx v8 for which that gene is expressed. From each CAFEH run we selected the components that are active in the GWAS (based on a threshold of 0.5) and active in one of the tissues that are enriched in CAD heritability (either one of three Artery tissues or the Liver). We also performed CAFEH jointly across all GTEx v8 tissues for all protein coding genes and selected the components that are active in either one of three Artery tissues or the Liver. We generated boxplots of the number of tissues that share each of the selected components in a 2×2 factorial design (CAD or GTEx tissue for each of the following tissues: Artery or Liver). The p-values in the figure represent p-values for the fixed effect term of linear mixed models of the form: number of tissues sharing component ∼ Group_of_component (CAD vs GTEx tissue)+ (1|gene).

In order to evaluate the effectiveness of CAFEH on prioritization of the target tissue in GWAS, we evaluated CAFEH’s performance in the same 19 highly powered GWA studies from the UK Biobank [37]. All GWA loci were evaluated for each study. For each individual run of CAFEH, the input was one of the GWAS loci and the cis-eQTL summary statistics of all genes that are significant cis-eGenes for the sentinel variant in the GWAS locus across all 49 GTEx v8 tissues. GTEx v8 LD was used as reference. We evaluated colocalization in any component or the top component (as defined previously) between the GWAS study and the GTEx tissues for each genome-wide significant locus and we counted the number of loci for which each tissue colocalizes with the corresponding GWAS (each locus could be counted for more than one tissues if it colocalizes with multiple tissues). If a GWAS locus sentinel variant was a significant eQTL for more than one genes, the locus was included in multiple CAFEH runs (one for each gene) and the counts of colocalizing tissues across runs for that locus were aggregated such that each tissue was counted one time if it colocalized with that locus for at least one tested gene. The results of the aggregate colocalization counts for each phenotype were compared to the results of partitioned LD score (LDSC) regression for the same phenotype using tissue specific genes from GTEx as previously described [38]. The number of loci colocalizing with partitioned LDSC-enriched vs non-enriched tissues was compared using a linear mixed model with each phenotype defined as a random effect. In addition, for more detailed visualization of the results, we ranked each tissue based on the number of loci colocalizing in each GTEx tissue and plotted the ranks in Supplementary Figure 18.

This analysis established that expected tissues based on partitioned LD score regression results and our prior knowledge of disease-specific pathogenesis can be identified and prioritized based on a colocalization approach for the majority of traits. We should note that our expectation is that partitioned LD score regression, by virtue of the fact that it leverages the whole GWAS signal as opposed to genome-wide significant loci may be more effective at this broad tissue prioritization. In contrast, our approach has the advantage of being able to identify putative target tissues at a locus resolution, therefore prioritizing tissues to be tested in downstream functional characterization of each locus. To demonstrate the effectiveness of CAFEH-S in identifying tissues in specific GWAS loci, we performed a case study using Coronary Artery Disease (CAD) as a complex trait. The choice of CAD was made based on the fact that it is a complex trait with a highly heritable component and with published high-powered GWA meta-analyses [25]. In addition, CAD has a multifactorial pathogenesis that involves multiple organ systems, thereby allowing for the possibility of different tissues being relevant in different loci. Lastly, since CAD was one of the first diseases to be studied in a GWAS approach, several downstream functional characterization studies have been undertaken for its significant loci, which provides gold-standard knowledge against which we can test the results of CAFEH-S.

To test CAFEH-S performance in CAD, we used a large-scale GWAS meta-analysis [25] and performed a literature review to select loci that either: (a) were in close proximity to a gene whose rare variants are known to predispose to atherosclerosis - the root cause of CAD - in a Mendelian fashion (LDLR, APOE, APOB, PCSK9, ANGPTL4, LPL, ABCG8, CETP), or (b) loci for which functional characterization followed by experimental validation studies have been performed linking the GWAS locus to a specific gene in a tissue or cell line [39-50]. For each of the above loci, we ran CAFEH-S jointly with the GWAS and the GTEx v8 eQTL summary statistics for the putative target gene across all 49 GTEx v8 tissues. We also ran COLOC pairwise between the GWAS locus and each tissue for the target gene with the same parameters as described previously. We then compared the number of loci that colocalize in the target tissue based on any component or the top component based on CAFEH-S and those that colocalized with the target tissue based on COLOC, stratified by whether the GWAS sentinel variant is a genome-wide significant eQTL for the target gene in the target tissue. We also plotted detailed results of colocalization based on CAFEH-S in Supplementary Figure 19.

## Data Availability

GTEx eQTL summary statistics are available in the GTEx portal (https://gtexportal.org/home/datasets). eQTLGen (https://eqtlgen.org/cis-eqtls.html) and MUTHER (http://www.muther.ac.uk/Data.html) summary statistics are available from the corresponding sources. GWAS summary statistics from the UK Biobank are publicly available for case control (https://www.leelabsg.org/resources) and continuous phenotypes (http://www.nealelab.is/uk-biobank). CAD GWAS summary statistics are available online (https://www.cardiomics.net/download-data).

## Acknowledgments

MA was supported by an NIH T32-HL007227 grant for this work.

## Software availability

CAFEH is available in github: https://github.com/karltayeb/cafeh

