## Supplementary Methods for "Redefining tissue specificity of genetic regulation of gene expression in the presence of allelic heterogeneity"

### CAFEH: Extended Methods

June 23, 2021

In this document we review variational inference and describe the variational approximation used in CAFEH. Then we derive the coordinate ascent updates for CAFEH-G and CAFEH-S. Finally, we describe how to use stochastic variational inference to improve speed of CAFEH-S optimization.

#### 1 Variational Inference Review

###### 1.0.1 Problem set up

Given a model $p(Y, \theta)$ where $Y$ are observed data and $\theta$ are latent variables, we want to compute the posterior distribution $p(\theta|Y)$. When the exact posterior distribution is intractable, we can approximate the posterior using variational inference.

In variational inference, we recast inference as an optimization problem. We posit a family of distributions $\mathcal{Q}$ over the latent variables in the model $\theta$ and find the member of that family that minimizes the KL-divergence to the true posterior.

$$q^*(\theta) = argmin_{q \in \mathcal{Q}} KL\left[q(\theta)||p(\theta|Y)\right] \tag{1}$$

When $p(\theta|Y) \in \mathcal{Q}$ this optimization yields the true posterior distribution. In practice, we choose $\mathcal{Q}$ so that we can efficiently optimize over the parameters of the family. Specifically it is often useful to choose a family of variational distributions that factorize over latent variables: $q(\theta) = \prod_i q(\theta_i)$.

We can solve this optimization by maximizing the Evidence Lower Bound (ELBO), which is a lower bound to the marginal data likelihood $p(Y|X) = \int_\theta p(Y, \theta|X) d\theta$

$$ELBO = \mathbb{E}_q\left[\ln p(Y, \theta|X)\right] + \mathbb{E}_q\left[\ln q(\theta)\right] \tag{2}$$

It can be shown that optimizing the ELBO with respect to the variational parameters is equivalent to minimizing the KL divergence in (1) [1].

The ELBO may be equivalently expressed as

$$ELBO = \mathbb{E}_q\left[p(Y|X, \theta)\right] - KL\left[q(\theta)||p(\theta)\right] \tag{3}$$

##### 1.0.2 Deriving updates

We want to derive the udpate for a variational factor $q(z)$. where $z$ is some subset of the latent variables in the model. Modifying the logic from [1] consider decomposing the ELBO

$$ELBO = \mathbb{E}_q(z)\left[\mathcal{L}\right] - \mathbb{E}_q(z)\left[\ln q(z)\right] + C \tag{4}$$

Where $\mathcal{L}$ are all terms of the ELBO that depend on $z$, and $q(z)$ is a density function which satisfies $\int q(z) = 1$. Using Lagrange multipliers to encode this constraint

$$\frac{d}{dq(z)}ELBO = \frac{d}{dq(z)}\left\{\mathbb{E}_{q(z)}\left[\mathbb{E}_{q(-z)}\left[\mathcal{L}\right]\right] - \mathbb{E}_{q(z)}\left[\ln q(z)\right] + \lambda\mathbb{E}_{q(z)}[1] - 1\right\} \tag{5}$$

$$= \mathbb{E}_{q(-z)}\left[\mathcal{L}\right] - \ln q(z) + \lambda \tag{6}$$

Setting the derivative equal to 0 we find

$$\ln q(z) = E_{q(-z)}\left[\mathcal{L}\right] + \lambda \tag{7}$$

Recognizing that $q(z)$ must integrate to one and that the normalizing factor does not depend on $z$

$$q^*(z) \propto exp\left\{\mathbb{E}_{q(-z)}\left[\mathcal{L}\right]\right\} \tag{8}$$

This suggests an approach for deriving our updates: compute $E_{q(-z)}\left[\mathcal{L}\right]$ and identify the parameters for $q(z)$ satisfying (8). Note that in general, identifying this distribution is not straight-forward. However, for a special class of models, of which CAFEH is a member, the coordinate-wise optima are exponential family distributions and their parameters can be computed analytically.

### 2 CAFEH-G

#### 2.1 Model

For clarity we restat the model. Let $Y$ an $N \times T$ matrix of measurements in $N$ individuals across $T$ phenotypes. Let $X$ be a $N \times G$ matrix of genotypes in $N$ individuals across $G$ SNPs. The CAFEH model is written as

$$
\begin{aligned}
Y_t &\sim (X\mathbf{b}_t, \tau_t^{-1} I) && (9)\\
\mathbf{b}_t &= \sum_{k=1}^{K} \phi_k w_{tk} s_{tk} && (10)\\
w_{tk}|\alpha_{tk} &\sim \mathcal{N}(0, \alpha_{tk}^{-1}) && (11)\\
s_{tk} &\sim \text{Bernoulli}(p_{0k}) && (12)\\
\phi_k &\sim \text{Categorical}(\pi_0) && (13)\\
\alpha_{tk} &\sim \Gamma(a_0, b_0) && (14)\\
\tau_t &\sim \Gamma(c_0, d_0) && (15)
\end{aligned}
$$

##### 2.2 Variational Approximation

Let $\theta = \{w_{tk}\} \cup \{s_{tk}\} \cup \{\phi_k\} \cup \{\alpha_{tk}\} \cup \{\tau_t\}$ denote the set of latent variables.

We select $\mathcal{Q}$ to factorize as follows:

$$
q^*(\theta) = \prod_k \prod_t q(w_{tk}|\phi_k, s_{tk}) q(s_{tk}) q(\alpha_{tk}) \prod_k q(\phi_k) \prod_t q(\tau_t) \quad (16)
$$

In particular we choose to a variational family that maintain dependence of $w_{tk}$ on $\phi_k$ and $s_{tk}$ so that we can accurately estimate effect sizes under different causal configurations. This is similar to the choice made in for the variational approximations chosen for SuSiE [3] and [2].

We optimize the ELBO via coordinate ascent, iteratively updating each $q(w|\phi, s)$, $q(\phi)$, $q(s)$, $q(\alpha)$ and $q(\tau)$, while holding the others fixed. Note, that while we have not specified a parametric form for the factors of the variational distribution, the model and factorization imply the optimal form of each variational factor:

$$
\begin{aligned}
q^*(s_{tk}) &\sim Bernoulli(\gamma_{tk})\\
q^*(\phi_k) &\sim Categorical(\pi_k)\\
q^*(\alpha_{tk}) &\sim \Gamma(a_{tk}, b_{tk})\\
q^*(\tau_t) &\sim \Gamma(c_t, d_t)\\
q^*(w_{tk}|\phi_k = i, s_{tk} = 1) &\sim \mathcal{N}(\mu_{tki}, \sigma_{tki}^2)
\end{aligned} \quad (17)
$$

$\{\mu, \sigma^2, \gamma, \pi, a, b, c, d\}$ (omitting subscripts) are *variational parameters* that we optimize over. We provide the full updates and their derivation below.

#### 2.3 Evidence Lower Bound (ELBO)

$$ELBO = \mathbb{E}_{q(\theta)}\left[\ln p(\mathbf{Y}|\theta)\right] - KL\left[q(\theta)||p(\theta)\right] \tag{18}$$

$$\begin{aligned}
&= \mathbb{E}_{q(\theta)}\left[\sum_t \ln \mathcal{N}(\mathbf{Y}_t|\mathbf{b}_t, \tau^{-1}I)\right] \\
&- \sum_{t,k} \mathbb{E}_{q(s_{tk},\alpha_{tk},\phi_k)}\left[KL\left[q(w_{tk}|s_{tk},\phi_k)||p(w_{tk}|\alpha_{tk})\right]\right] \\
&- \sum_{t,k} KL\left[q(s_{tk})||p(s_{tk})\right] - \sum_{t,k} KL\left[q(\alpha_{tk})||p(\alpha_{tk})\right] \\
&- \sum_{k} KL\left[q(\phi_k)||p(\phi_k)\right] - \sum_t KL\left[q(\tau_t||p(\tau_t)\right]
\end{aligned} \tag{19}$$

##### 2.3.1 Expected conditional

$$\begin{aligned}
&\mathbb{E}_{q(\theta)}\left[\ln \mathcal{N}(\mathbf{Y}_t|\mathbf{X}\mathbf{b}_t, \tau^{-1}I)\right] = \\
&\quad -\frac{M}{2}\ln 2\pi + \frac{M}{2}\left\langle \ln \tau_t \right\rangle - \frac{\langle \tau_t \rangle}{2}\left[\mathbf{Y}_t^T\mathbf{Y}_t - 2\mathbf{Y}_t^T\left\langle \mathbf{X}\mathbf{b}_t\right\rangle - \left\langle \mathbf{b}_t^T\mathbf{X}^T\mathbf{X}\mathbf{b}_t\right\rangle\right]
\end{aligned} \tag{20}$$

The expectation of $\mathbf{b}_t$ is

$$\langle \mathbf{b}_t \rangle = \sum_k (\pi_k \circ \mu_{tk})\gamma_{tk} \tag{21}$$

Letting $d_i = e_i^T\mathbf{X}^T\mathbf{X}e_i$ and $\langle \mathbf{b}_{tk}\rangle = (\pi_k \circ \mu_{tk})\gamma_{tk}$ and noting $s_{tk}^2 = s_{tk}$ we can get a nice expression for the quadratic term

$$\left\langle \mathbf{b}_t^T\mathbf{X}^T\mathbf{X}\mathbf{b}_t\right\rangle = \left\langle \left(\sum_k \phi_k w_{tk} s_{tk}\right)^T \mathbf{X}^T\mathbf{X}\left(\sum_k \phi_k w_{tk} s_{tk}\right)\right\rangle \tag{22}$$

$$= \sum_k \left\langle w_{tk}^2 s_{tk} d_{\phi_k}\right\rangle + \sum_{k\neq j}\left\langle w_{tk}s_{tk}\phi_k^T\right\rangle \mathbf{X}^T\mathbf{X}\left\langle \phi_j w_{tj} s_{tj}\right\rangle \tag{23}$$

$$= \sum_{k,i}(\mu_{tki}^2 + \sigma_{tki}^2)\gamma_{tk}\pi_{ki}d_i + \langle \mathbf{b}_t\rangle^T \mathbf{X}^T\mathbf{X}\langle \mathbf{b}_t\rangle - \sum_k ||\mathbf{X}\langle \mathbf{b}_{tk}\rangle||^2 \tag{24}$$

##### 2.3.2 KL computations

To compute the ELBO and coordinate ascent updates, we need to compute $\mathbb{E}\left[KL\left[q(w|\phi,s)||p(w|\alpha)\right]\right]$, where expectations are taken over $q(\alpha)$, $q(s_{tk})$ and/or $q(\phi_k)$ depending on the setting. $s$ and $\phi$ appear linearly, while $\alpha$ does not. Here we write the expectation of the KL divergence w.r.t $\alpha$ in terms of the the KL of the expectation plus a positive correction.

$$\left\langle KL\left[\mathcal{N}(\mu, \sigma^2)\,||\mathcal{N}(0, \alpha^{-1})\right.\right. \tag{25}$$

$$= \left\langle \frac{1}{2}\left[\alpha\mu^2 + \sigma^2\alpha - 1 - \ln\sigma^2 - \ln\alpha\right]\right\rangle \tag{26}$$

$$= \frac{1}{2}\left[\langle\alpha\rangle\mu^2 + \sigma^2\langle\alpha\rangle - 1 - \ln\sigma^2 - \langle\ln\alpha\rangle\right] \tag{27}$$

$$= \frac{1}{2}\left[\langle\alpha\rangle\mu^2 + \sigma^2\langle\alpha\rangle - 1 - \ln\sigma^2 - \ln\langle\alpha\rangle\right] + \frac{1}{2}(\ln\langle\alpha\rangle - \langle\ln\alpha\rangle) \tag{28}$$

$$= KL\left[\mathcal{N}(\mu, \sigma^2)||\mathcal{N}(0, \langle\alpha\rangle^{-1})\right] + \frac{1}{2}(\ln\langle\alpha\rangle - \langle\ln\alpha\rangle) \tag{29}$$

##### 2.3.3 Residualized likelihood

As we write our variational updates it will be useful to define $r_{tk} = Y_t - X\mathbf{b}_t + X\mathbf{b}_{tk}$ where $\mathbf{b}_{tk} = \phi_k w_{tk} s_{tk}$. That is, $r_{tk}$ is the residual with all but the $k$-th component removed. The conditional likelihood may be written

$$\mathcal{N}(Y_t|X\mathbf{b}_t, \tau_t^{-1}) = \mathcal{N}(r_{tk}|X\mathbf{b}_{tk}, \tau_t^{-1}) \tag{30}$$

Then, when considering updates for a particular component $k$, we can write the ELBO as

$$ELBO = \mathbb{E}_{q(\theta)}\left[\sum_t -\frac{\tau_t}{2}\left[-2r_{tk}^T\mathbf{X}\mathbf{b}_{tk} + \mathbf{b}_{tk}^T\mathbf{X}^T\mathbf{X}\mathbf{b}_{tk}\right]\right] - KL\left[q(\theta)||p(\theta)\right] \tag{31}$$

#### 2.4 Coordinate Ascent updates

##### 2.4.1 Update for $q^*(w_{tk}|\phi_k = i, s_{tk} = 1$

Where $\mathbf{x}_i$ is the $i$th column of $X$, the genotypes at SNP $i$.

$$q^*(w_{tk}|s_{tk} = 1, \phi_k = i) \tag{32}$$

$$\propto exp\left\{\left\langle\ln\mathcal{N}(r_{tk}|w_{tk}\mathbf{x}_i, \tau_t^{-1}\mathbf{I})\right\rangle + \langle\ln p(w_{tk}|\alpha_{tk})\rangle\right\} \tag{33}$$

$$\propto exp\left\{\ \frac{\langle\tau_t\rangle}{2}\left(-2\langle r_{tk}\rangle^T\mathbf{x}_i w_{tk} + d_i w_{tk}^2\right) + \frac{\langle\alpha_{tk}\rangle}{2}(w_{tk}^2)\right\} \tag{34}$$

Completing the square we find

$$\sigma_{tki}^2 = (d_i\langle\tau_t\rangle + \langle\alpha_{tk}\rangle)^{-1} \tag{35}$$

$$\mu_{tki} = \sigma_{tki}^2\langle\tau_t\rangle\langle r_{tk}\rangle^T\mathbf{x}_i \tag{36}$$

$$q^*(w_{tk}|\phi_k = i, s_{tk} = 1) = \mathcal{N}(w_{tk}|\mu_{tki}, \sigma_{tki}^2) \tag{37}$$

###### 2.4.2 Update for $q^*(w_{tk}|\phi_k, s_{tk}=0)$

$$
\begin{aligned}
& q^*(w_{tk}|s_{tk}=1, \phi_k = i) \\
& \qquad \propto exp\left\{\left\langle \ln \mathcal{N}(r_{tk}|0, \tau_t^{-1}\mathbf{I})\right\rangle + \left\langle \ln p(w_{tk}|\alpha_{tk})\right\rangle\right\} \\
& \qquad \propto exp\left\{\frac{\langle \alpha_{tk}\rangle}{2}(w_{tk}^2)\right\}
\end{aligned} \tag{38}
$$

$$
q^*(w_{tk}|s_{tk}=0, \phi_k = i) = \mathcal{N}(w+tk|0, \langle \alpha_{tk}\rangle^{-1}) \;\; \forall i \in \{1, \dots, N\} \tag{39}
$$

###### 2.4.3 Update for $q^*(s_{tk})$

$$
\begin{aligned}
q^*(s_{tk}) \propto exp\{ & \left\langle \ln \mathcal{N}(r_{tk}|\mathbf{X}\phi_k w_{tk}, \tau_t^{-1}\mathbf{I})\right\rangle \mathbb{1}(s_{tk}=1) \\
& + \langle KL\left[q(w_{tk}, \alpha_{tk}|s_{tk}=1, \phi_k)||p(w_{tk}, \alpha_{tk})\right]\rangle \mathbb{1}(s_{tk}=1) \\
& + \ln p_{0k}\mathbb{1}(s_{tk}=1) \\
& \left\langle \ln \mathcal{N}(r_{tk}|0, \tau_t^{-1}\mathbf{I})\right\rangle \mathbb{1}(s_{tk}=0) \\
& + \langle KL\left[q(w_{tk}, \alpha_{tk}|s_{tk}=0, \phi_k)||p(w_{tk}, \alpha_{tk})\right]\rangle \mathbb{1}(s_{tk}=0) \\
& + \ln(1-p_{0k})\mathbb{1}(s_{tk}=0)
\end{aligned} \tag{40}
$$

Grouping terms where $s_{tk}=1$ and $s_{tk}=0$ we can write

$$
q^*(s_{tk}) \propto exp\left\{(a + \ln p_{0k})\mathbb{1}(s_{tk}=1) + (b + \ln(1-p_{0k}))\mathbb{1}(s_{tk=0})\right\} \tag{41}
$$

$$
\begin{aligned}
a = & -\frac{\langle \tau_t\rangle}{2}\left[-2\left\langle r_{tk}\right\rangle^T \mathbf{X}(\pi_k \circ \mu_{tk}) + \sum_i (\mu_{tki}^2 + \sigma_{tki}^2)\pi_{ki}\right] \\
& - \sum_i \pi_{ki}\left\langle KL\left[q(w_{tk}, \alpha_{tk}|s_{tk}=1, \phi_k = i)||p(w_{tk}, \alpha_{tk}\right]\right\rangle
\end{aligned} \tag{42}
$$

$$
b = -\left\langle KL\left[q(w_{tk}, \alpha_{tk}|s_{tk}=0)||p(w_{tk}, \alpha_{tk}\right]\right\rangle = -\frac{1}{2}(\ln\langle\alpha\rangle - \langle \ln \alpha\rangle) \tag{43}
$$

Setting $\gamma_{tk} = \frac{e^a p_{0k}}{e^a p_{0k} + e^b(1-p_{0k})}$

$$
q^*(s_{tk}) = \text{Bernoulli}(s_{tk}|\gamma_{tk}) \tag{44}
$$

###### 2.4.4 Update for $q^*(\alpha_{tk})$

$$
\begin{aligned}
q^*(\alpha_{tk}) &\propto exp\left\{\left\langle \ln\mathcal{N}(w_{tk}|0,\alpha_{tk}^{-1})\ln p(\alpha_{tk})\right\rangle\right\} \\
&\propto exp\left\{\frac{1}{2}\ln\alpha_{tk} - \frac{\alpha_{tk}}{2}\left\langle w_{tk}^2\right\rangle + (a_0-1)\ln\alpha_{tk} - b_0\alpha_{tk}\right\} \\
&\propto exp\left\{\left(a_0+\frac{1}{2}-1\right)\ln\alpha_{tk} - \left(b_0+\frac{\left\langle w_{tk}^2\right\rangle}{2}\right)\alpha_{tk}\right\} \\
&\propto exp\left\{\left(a_0+\frac{1}{2}-1\right)\ln\alpha_{tk} - \left(b_0+\frac{\sum_i \pi_{ki}(\mu_{tki}^2+\sigma_{tki})^2}{2}\right)\alpha_{tk}\right\}
\end{aligned}
\tag{45}
$$

Let $a = a_0 + \frac{1}{2}$ and $b = b_0 + \frac{\sum_i \pi_{ki}(\mu_{tki}^2+\sigma_{tki})^2}{2}$

$$
q^*(\alpha_{tk}) = \Gamma(\alpha_{tk}|a,b) \tag{46}
$$

###### 2.4.5 Update for $q^*(\phi_k)$

$$
q^*(\phi_k) \propto \sum_i \rho_{ki} 1(\phi_k = i) \tag{47}
$$

$$
\begin{aligned}
\rho_{ki} = &\left\langle \ln\mathcal{N}(r_{tk}|s_{tk}w_{tk}\mathbf{x}_i, \tau^{-1}I\right\rangle \\
&- \langle KL[q(w_{tk},\alpha_{tk}|\phi_k = i)\;||\;p(w_{tk}|\alpha_{tk})]\rangle + \ln\pi_{0ki}
\end{aligned}
\tag{48}
$$

$$
\begin{aligned}
\rho_{ki} = &-\frac{\langle\tau_t\rangle}{2}\left[-2\left\langle r_{tk}\right\rangle^T\mathbf{x}_i\mu_{tk}\gamma_{tk} + \gamma_{tk}(\mu_{tki}^2+\sigma_{tki}^2)\right] \\
&- \langle KL[q(w_{tk},\alpha_{tk}|s_{tk}=1,\phi_k = i)\;||\;p(w_{tk}|\alpha_{tk})]\rangle\,\gamma_{tk} \\
&- \langle KL[q(w_{tk},\alpha_{tk}|s_{tk}=0,\phi_k = i)\;||\;p(w_{tk}|\alpha_{tk})]\rangle\,(1-\gamma_{tk}) + \ln\pi_{0ki}
\end{aligned}
\tag{49}
$$

Then

$$
\pi_{ki} = \frac{e^{\rho_i}}{\sum_i e^{\rho_{ik}}} \tag{50}
$$

###### 2.4.6 Update for $q^*(\tau_t)$

$$
\begin{aligned}
\ln q^*(\tau_t) &\propto \left\langle \mathcal{N}(\hat{\beta}_t|\mathbf{X}\mathbf{b}_t,\tau_t^{-1}I) + \ln p(\tau_t)\right\rangle \\
&\propto \frac{1}{2}\ln\tau_t - \frac{\tau_t}{2}\left\langle(\hat{\beta}_t - \mathbf{X}\mathbf{b}_t)^T(\hat{\beta}_t - \mathbf{X}\mathbf{b}_t)\right\rangle + (c_0-1)\ln\tau_t - d_0\tau_t
\end{aligned}
\tag{51}
$$

Let $c = c_0 + \frac{1}{2}$ and $d = d_0 + \frac{\left\langle(\hat{\beta}_t - \mathbf{X}\mathbf{b}_t)^T(\hat{\beta}_t - \mathbf{X}\mathbf{b}_t)\right\rangle}{2}$

$$
q^*(\tau_t) = \Gamma(\tau_t|c,d) \tag{52}
$$

### 3 CAFEH-S model

CAFEH-S has an identical prior on the effect sizes $\mathbf{b}_t$ as CAFEH-G, however the likelihood is written in terms of summary statistics using the RSS likelihood [4]. $\hat{\beta}_t$ are the vector of effect sizes for marginal linear regression of $G$ SNPs in phenotype $t$. $R$ is an LD matrix containing the pairwise correlation of SNPs. $S$ is a diagonal matrix where $S_{ii}^2 = \beta^2/n_{ti} + \hat{s}^2 + ti$. $n_{ti}$ and $\hat{s}_{ti}$ are the sample size and standard errors for the corresponding tests.

$$\hat{\beta}_t \sim (SRS^{-1}\mathbf{b}_t, SRS) \tag{53}$$

$$\mathbf{b}_t = \sum_{k=1}^{K} \phi_k w_{tk} s_{tk} \tag{54}$$

$$w_{tk}|\alpha_{tk} \sim \mathcal{N}(0, \alpha_{tk}^{-1}) \tag{55}$$

$$s_{tk} \sim \text{Bernoulli}(p_{0k}) \tag{56}$$

$$\phi_k \sim \text{Categorical}(\pi_0) \tag{57}$$

$$\alpha_{tk} \sim \Gamma(a_0, b_0) \tag{58}$$

#### 3.1 Evidence Lower Bound (ELBO)

We write the ELBO, lumping terms that are constant w.r.t the variational parameters into a constant $C$. Letting

$$D = S^{-1}RS^{-1}$$

$$ELBO = \mathbb{E}_q\left[\sum_t ln\mathcal{N}(\hat{\beta}_t|SRS^{-1}\mathbf{b}_t, SRS)\right] - KL\left[q||p\right] \tag{59}$$

$$= \mathbb{E}_q\left[\sum_t -\frac{1}{2}\left(-2\hat{\beta}_t^T S^{-2}\mathbf{b}_t + \mathbf{b}_t^T D\mathbf{b}_t\right)\right] - KL\left[q||p\right] + C \tag{60}$$

##### 3.1.1 Residualized likelihood

Our coordinate ascent updates are performed by updating one component while holding all other components and fixed. It will be convenient to rewrite the likelihood in terms of the residual with all but one component removed

$$\mathbf{b}_{tk} = w_{tk}s_k\phi_k \tag{61}$$

$$\mathbf{b}_{-tk} = \sum_{j\neq k}\mathbf{b}_{tj} \tag{62}$$

$$r_{tk} = \hat{\beta}_t - SRS^{-1}\mathbf{b}_{-tk} \tag{63}$$

So that

$$\mathcal{N}(\hat{\beta}_t|SRS^{-1}\mathbf{b}_t, SRS) = \mathcal{N}(r_{tk}|SRS^{-1}\mathbf{b}_{tk}, SRS) \tag{64}$$

Notice that the term $r_{tk}^T(SRS)^{-1}r_{tk}$ does not depend on component $k$. For the purpose of optimization of the variational parameters of component $k$ we may write the ELBO

$$ELBO = \mathbb{E}_q\left[\sum_t -\frac{1}{2}\left(-2r_{tk}^T S^{-2}\mathbf{b}_t + \mathbf{b}_t^T D\mathbf{b}_t\right)\right] - KL\left[q||p\right] + C \tag{65}$$

#### 3.2 Coordinate Ascent updates

##### 3.2.1 Update for $q^*(w_{tk}|\phi_k, s_{tk} = 1)$

With $d_i = D_{ii}$

$$\begin{aligned} &q^*(w_{tk}|s_{tk} = 1, \phi_k = i) \propto \\ &\qquad exp\left\{\left\langle \ln\mathcal{N}(r_{tk}|SRS^{-1}\mathbf{b}_{tk}, SRS) + \ln\mathcal{N}(w_{tk}|0, \alpha_{tk})\right\rangle\right\} \\ &\qquad exp\left\{-\frac{1}{2}\left[-2\left\langle r_{tk}\right\rangle^T S^{-2}e_i w_{tk} + d_i w_{tk}^2 + \left\langle \alpha_{tk}\right\rangle w_{tk}^2\right]\right\} \end{aligned} \tag{66}$$

Completing the square we arrive at

$$\begin{aligned} \sigma_{tki}^2 &= (d_i + \langle\alpha\rangle)^{-1} \\ \mu_{tki} &= \sigma_{tki}^2 \left\langle r_{tk}\right\rangle^T S^{-2}e_i \\ q^*(w_{tk}|\phi_k = i, s_{tk} = 1) &= \mathcal{N}(w_{tk}|\mu_{tki}, \sigma_{tki}^2) \end{aligned} \tag{67}$$

##### 3.2.2 Update for $q^*(w_{tk}|\phi_k, s_{tk} = 0)$

$$q^*(w_{tk}|s_{tk} = 0, \phi_k = i) \propto exp\left\{-\frac{1}{2}\left\langle\alpha_{tk}\right\rangle w_{tk}^2\right\} \tag{68}$$

It follows that

$$q^*(w_{tk}|\phi_k, s_{tk} = 0) = \mathcal{N}(w_{tk}|0, \left\langle\alpha_{tk}\right\rangle^{-1}) \tag{69}$$

##### 3.2.3 Update for $q^*(s_{tk})$

We group terms of the ELBO where $s_{tk} = 1$:

$$\begin{aligned} a = \mathbb{E}_{q|s_{tk}=1}&\left[\log\mathcal{N}(r_{tk}|SRS^{-1}b_{tk}, SRS)\right] \\ &+\mathbb{E}_{q(w_{tk},\phi_k,s_{tk}=1)}\left[\log p(w_{tk}|\alpha_{tk})\right] \\ +\mathbb{E}_{q(\phi_k)}&\left[H(q(w_{tk}|s_{tk} = 1, \phi_k)\right] + \log p_{0k} + C \end{aligned} \tag{70}$$

Evaluates to

$$a = -\frac{1}{2}\left(-2\left\langle r_{tk}\right\rangle^T S^{-2}(\pi_k \circ \mu_{tk}) + \sum_i (\mu_{tki}^2 + \sigma_{tki}^2) d_i \pi_{ki}\right) \\ +\mathbb{E}_{q(w_{tk},\phi_k,s_{tk}=1)}\left[\log p(w_{tk}|\alpha_{tk})\right] \\ +\mathbb{E}_{q(\phi_k)}\left[H(q(w_{tk}|s_{tk}=1,\phi_k)\right] + \log p_{0k} + C \tag{71}$$

And $s_{tk} = 0$:

$$b = \mathbb{E}_{q|s_{tk}=0}\left[\log \mathcal{N}(r_{tk}|SRS^{-1}b, SRS)\right] \\ +\mathbb{E}_{q(w_{tk},\phi_k,s_{tk}=0)}\left[\log p(w_{tk}|\alpha_{tk})\right] + \\ \mathbb{E}_{q(\phi_k)}\left[H(q(w_{tk}|s_{tk}=0,\phi_k)\right] + \log(1-p_{0k}) + C \tag{72}$$

Evaluates to

$$b = 0 \\ +\mathbb{E}_{q(w_{tk},\phi_k,s_{tk}=0)}\left[\log p(w_{tk}|\alpha_{tk})\right] \\ +\mathbb{E}_{q(\phi_k)}\left[H(q(w_{tk}|s_{tk}=0,\phi_k)\right] + \\ \log(1-p_{0k}) + C \tag{73}$$

$$q^*(s_{tk}) \propto exp\left\{a1(s_{tk}=1) + b1(s_{tk}=0)\right\} \implies \gamma_{tk} = \frac{e^a}{e^a + e^b} \tag{74}$$

###### 3.2.4 Update for $q^*(\phi_k)$

Grouping terms where $\phi_k = i$

$$a_i = \mathbb{E}_{q|\phi_k=i}\left[\log \mathcal{N}(r_{tk}|SRS^{-1}b_{tk}, SRS)\right] \\ +\mathbb{E}_{q(w_{tk},s_{tk}|\phi_k=i)}\left[p(w_{tk}|\alpha_{tk})\right] \\ +\mathbb{E}_{q(s_{tk})}\left[H(q(w_{tk}|s_{tk},\phi_k=i)\right] \tag{75}$$

$$a_i = -\frac{1}{2}\left[-2\left\langle r_{tk}\right\rangle^T S^{-2} e_i \mu_{tki}\gamma_{tk} + \gamma_{tk}(\mu_{tk}^2 + \sigma_{tki}^2) d_i\right] \\ +\mathbb{E}_{q(w_{tk},s_{tk}|\phi_k=i)}\left[p(w_{tk}|\alpha_{tk})\right] \\ +\mathbb{E}_{q(\phi_k)}\left[H(q(w_{tk}|s_{tk},\phi_k=i)\right] \tag{76}$$

$$q^*(s_{tk}) \propto exp\left\{\sum_i a_i 1(\phi_k = i)\right\} \implies \pi_{ki} = \frac{e^{a_i}}{\sum_i e^{a_i}} \tag{77}$$

#### 3.3 Stochastic Variational Inference

##### 3.3.1 Monte-Carlo estimate of the ELBO

Recall the ELBO for CAFEH-S

$$ELBO = \mathbb{E}_q \left[ \sum_t -\frac{1}{2} \left( -2 r_{tk}^T S^{-2} \mathbf{b}_t + \mathbf{b}_t^T D \mathbf{b}_t \right) \right] - KL\left[q||p\right] + C \tag{78}$$

The CAFEH-S updates, (equivalently, evaluating the gradient of the ELBO), require the repeated evaluation of $\langle r_{tk} \rangle = \hat{\beta}_t - SRS^{-1} \langle \mathbf{b}_{-tk} \rangle$. This involves a matrix-vector multiplication that grows with the number of SNPs, and causes CAFEH-S to be slow to run with a large number of variants.

We propose using a Monte-Carlo estimate for the expectation over $q(\phi)$. Rather than averaging over all SNPs, and incurring the expensive matrix-vector multiplication, we sample SNPs. We write $\mathbf{b}_{tk}(\phi_k)$ to emphasize the dependence of $\mathbf{b}_{tk}$ on $\phi_k$.

$$\mathbb{E}_{q(\phi_k)} \left[ \mathbb{E}_{q(-\phi_k)} \mathbf{b}_{tk}(\phi_k) \right] \approx \frac{1}{L} \sum_{l=1}^{L} \mathbb{E}_{q(-\phi_k)} \mathbf{b}_{tk}(z_k^{(l)}) = \tilde{\mathbf{b}}_{tk} \tag{79}$$

Where $z_k^{(1)}, \ldots, z_k^{(L)}$ are iid samples from $Categorical(\pi_k)$, the current setting of $q(\phi_k)$. This approximation yields a noisy but unbiased estimate of the ELBO, satisfying the core requirement for performing stochastic optimization.

Importantly for moderate choice of $L$, $LK << G$. Thus, $\tilde{\mathbf{b}}_t$ is sparse and $SRS^{-1}\tilde{\mathbf{b}}_{tk}$ can be computed quickly.

##### 3.3.2 Stochastic Variational Inference

For models where all the complete conditionals are an exponential family, coordinate ascent on stochastic estimates of the ELBO is stochastic gradient ascent (in the natural parameter space) [cite]. In short, we can use the same updates as above, replacing expectations over $q(\phi_k)$ with their Monte-Carlo estimate, to compute $\hat{\lambda}$ an intermediate estimate of our variational parameter $\lambda$. We update our estimate of $\lambda$ as a weighted average of our old estimate and the intermediate estimate

$$\lambda^{(t+1)} = (1 - \rho_t)\lambda_t + \rho_t \hat{\lambda}_t \tag{80}$$

Where $t$ indicates iteration, and $\rho_t$ are weights. When the sequence $(\rho_t)_{t=1}^{\infty}$ satisfy the Robbins Monro conditions $\sum \rho_t = \infty$ and $\sum \rho_t^2 < \infty$, the stochastic optimization is guaranteed to converge to a local optimum.

We note that for well behaved causal components, where $q(\phi_k)$ places most of its mass on a set of tightly linked SNPs, the Monte-Carlo estimate will be very close to the true expectation.
