## Supplementary Figure for "Redefining tissue specificity of genetic regulation of gene expression in the presence of allelic heterogeneity"

### Slide 1
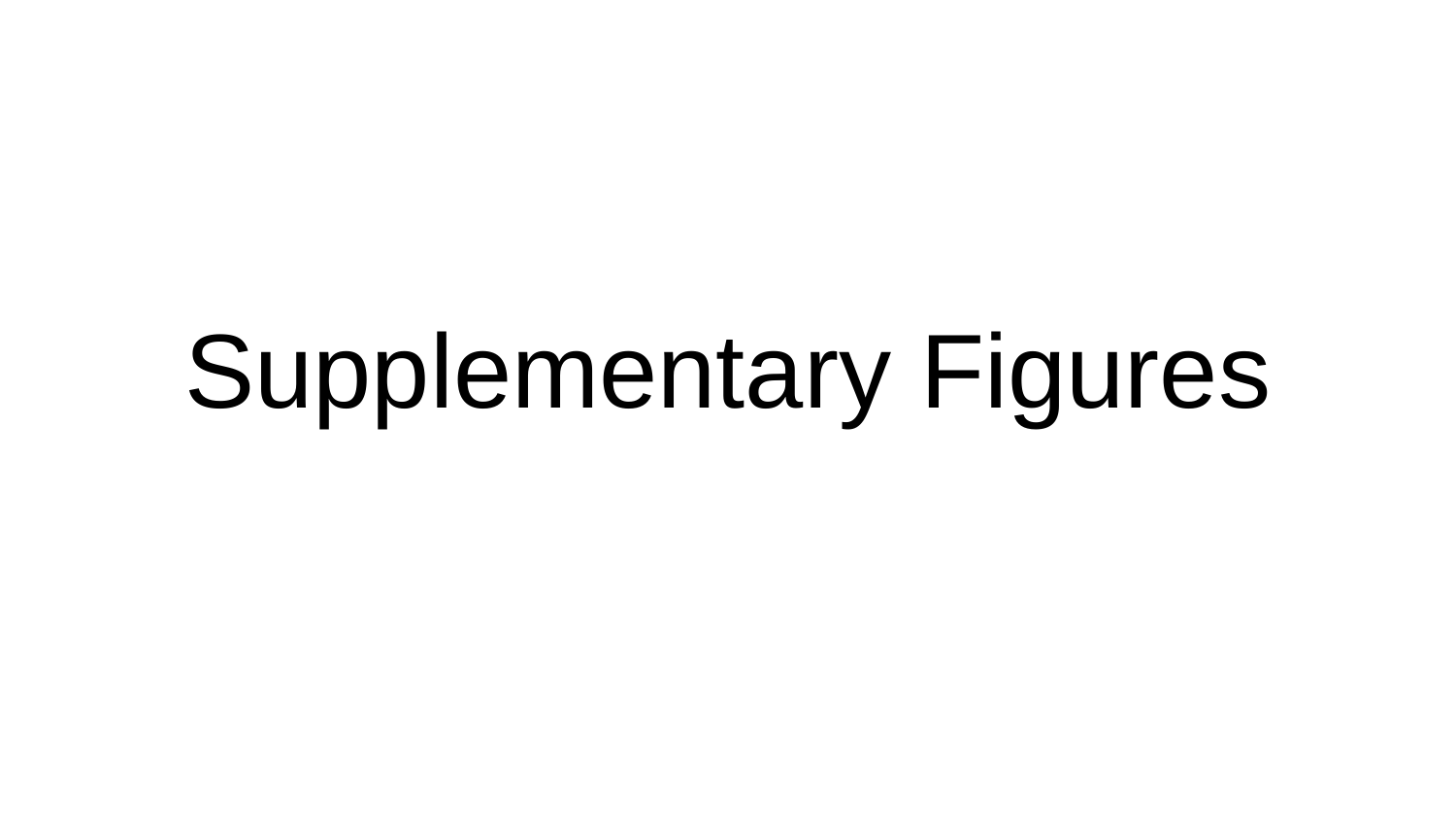

#### Supplementary Figures

### Slide 2
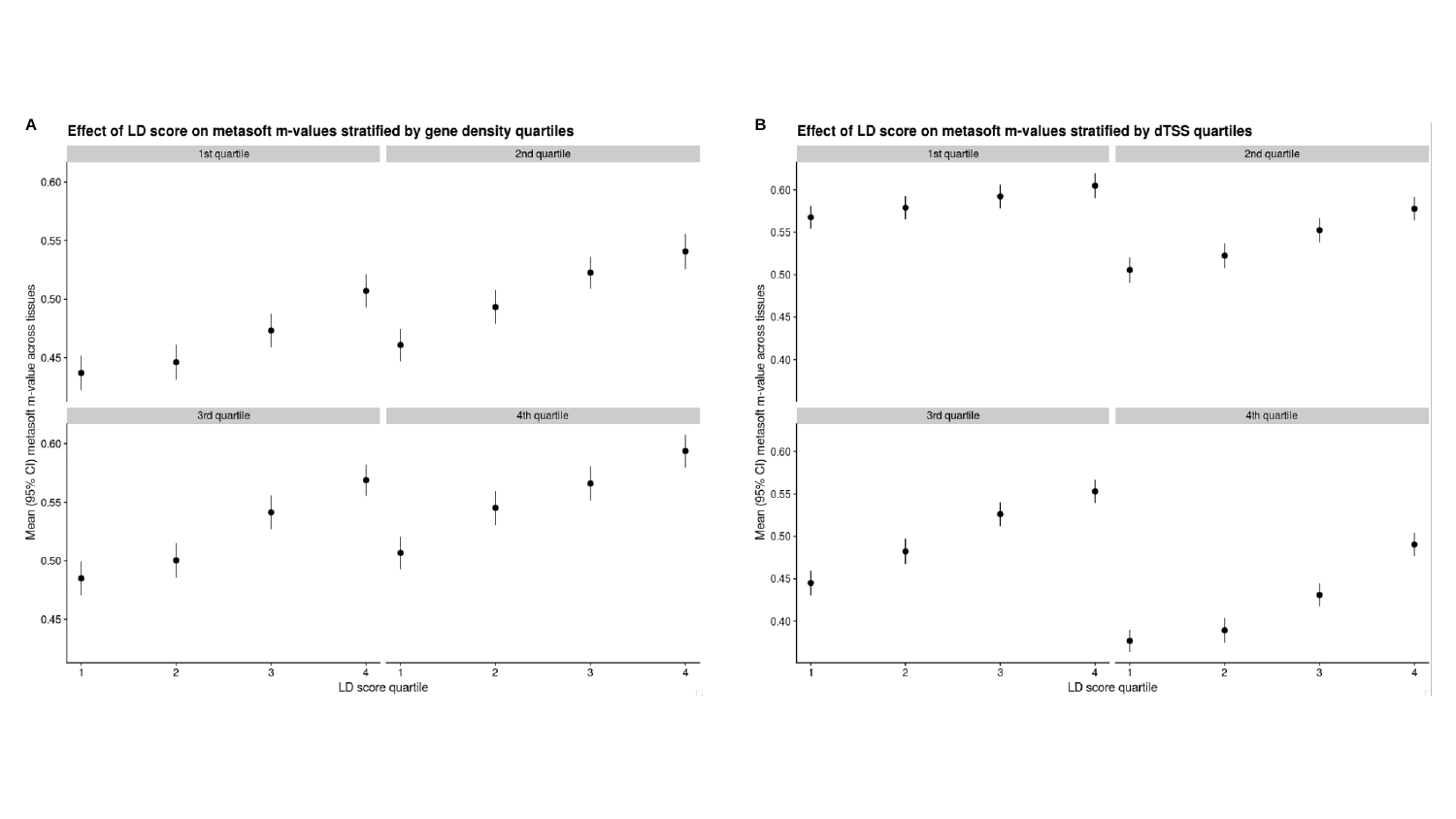

A
B

### Slide 3
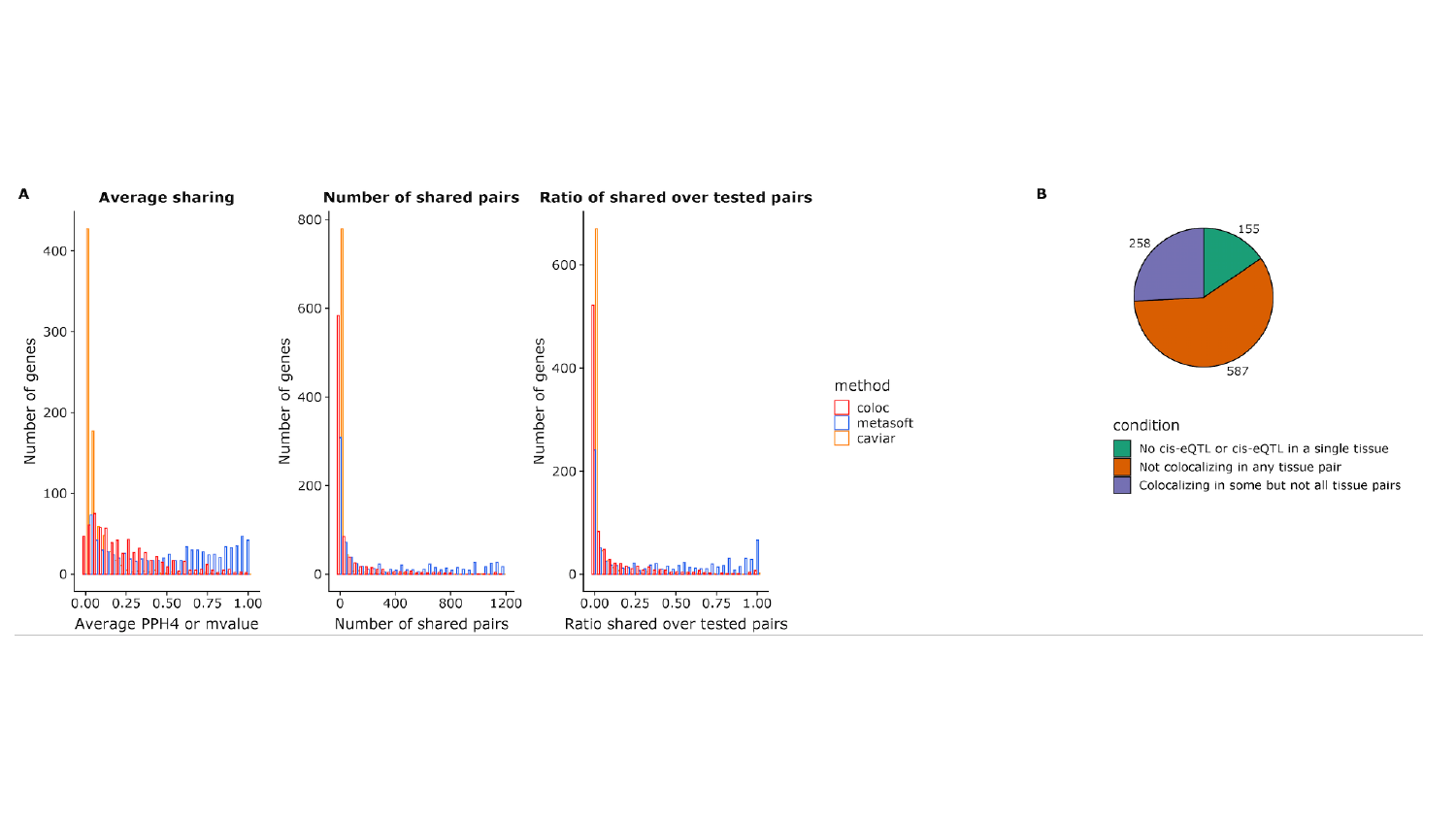

### Slide 4
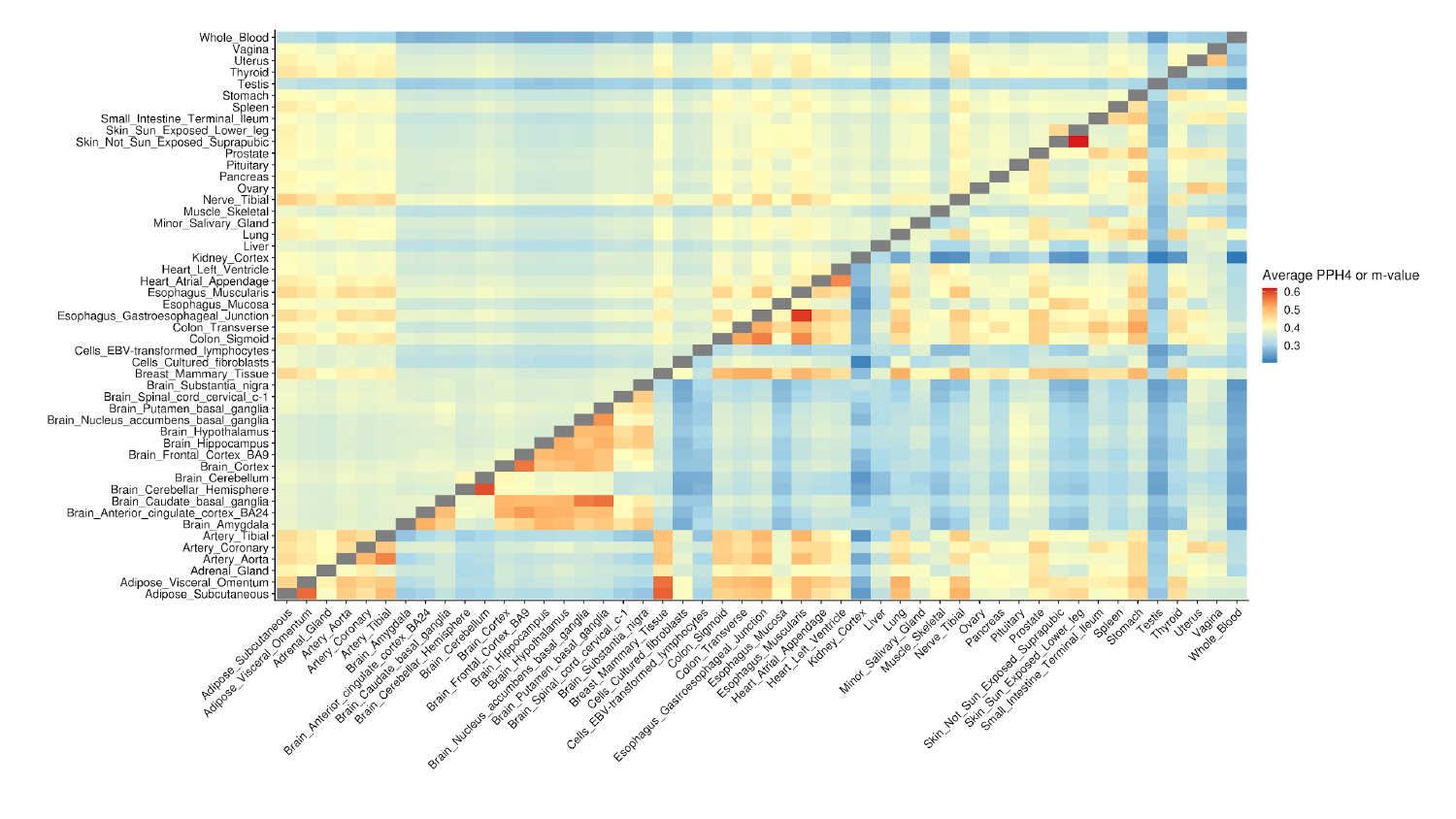

### Slide 5
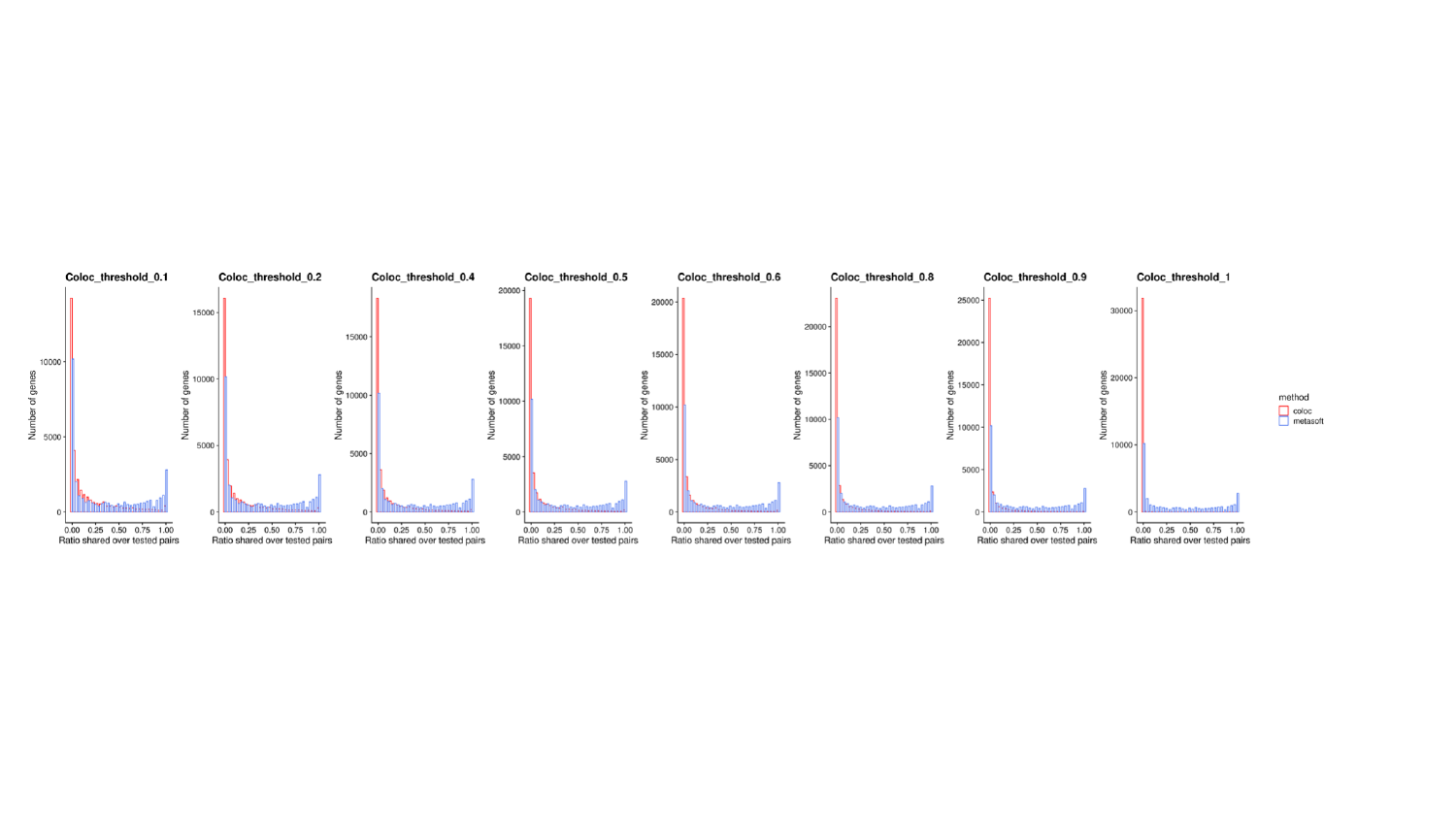

### Slide 6
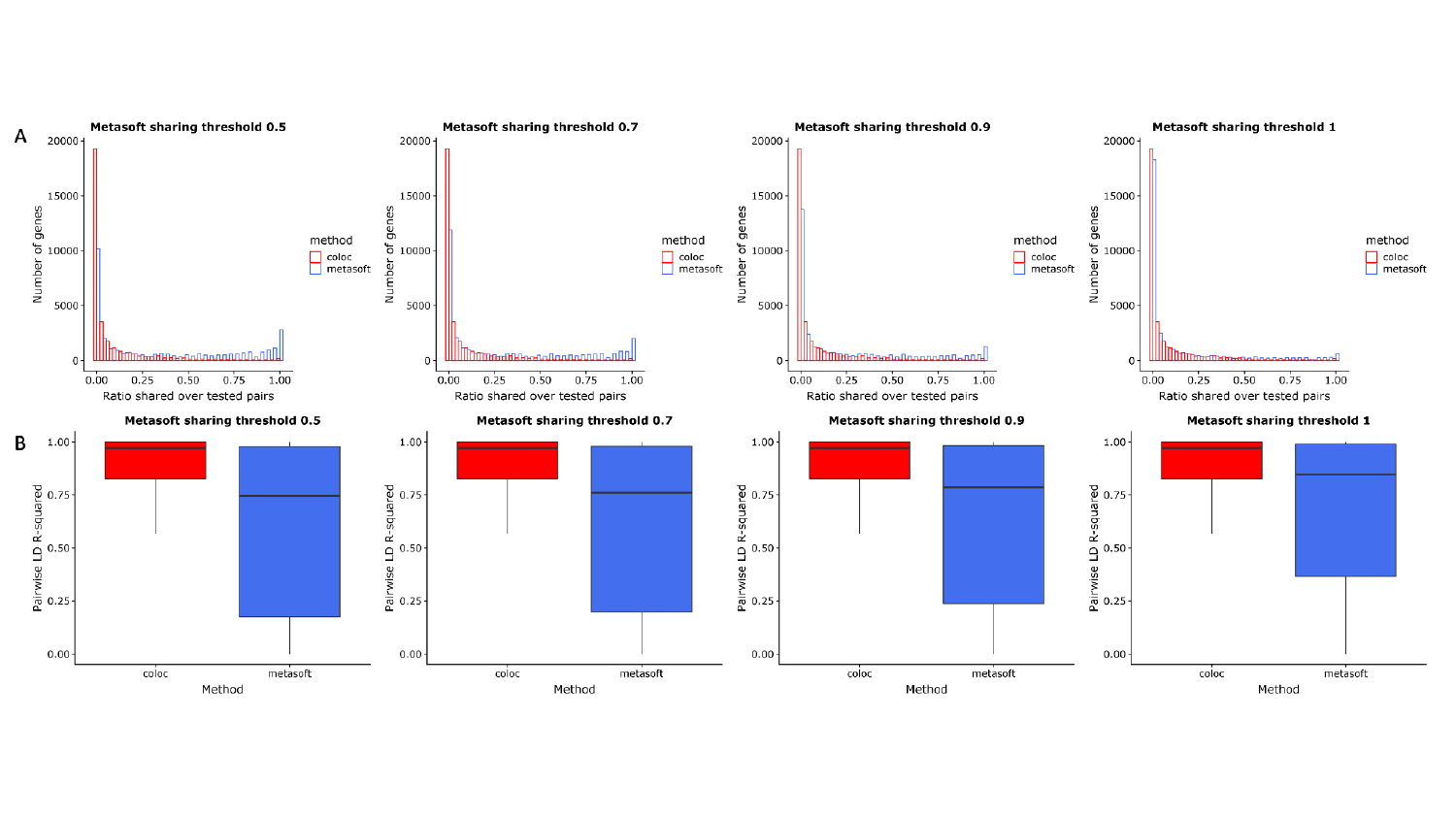

### Slide 7
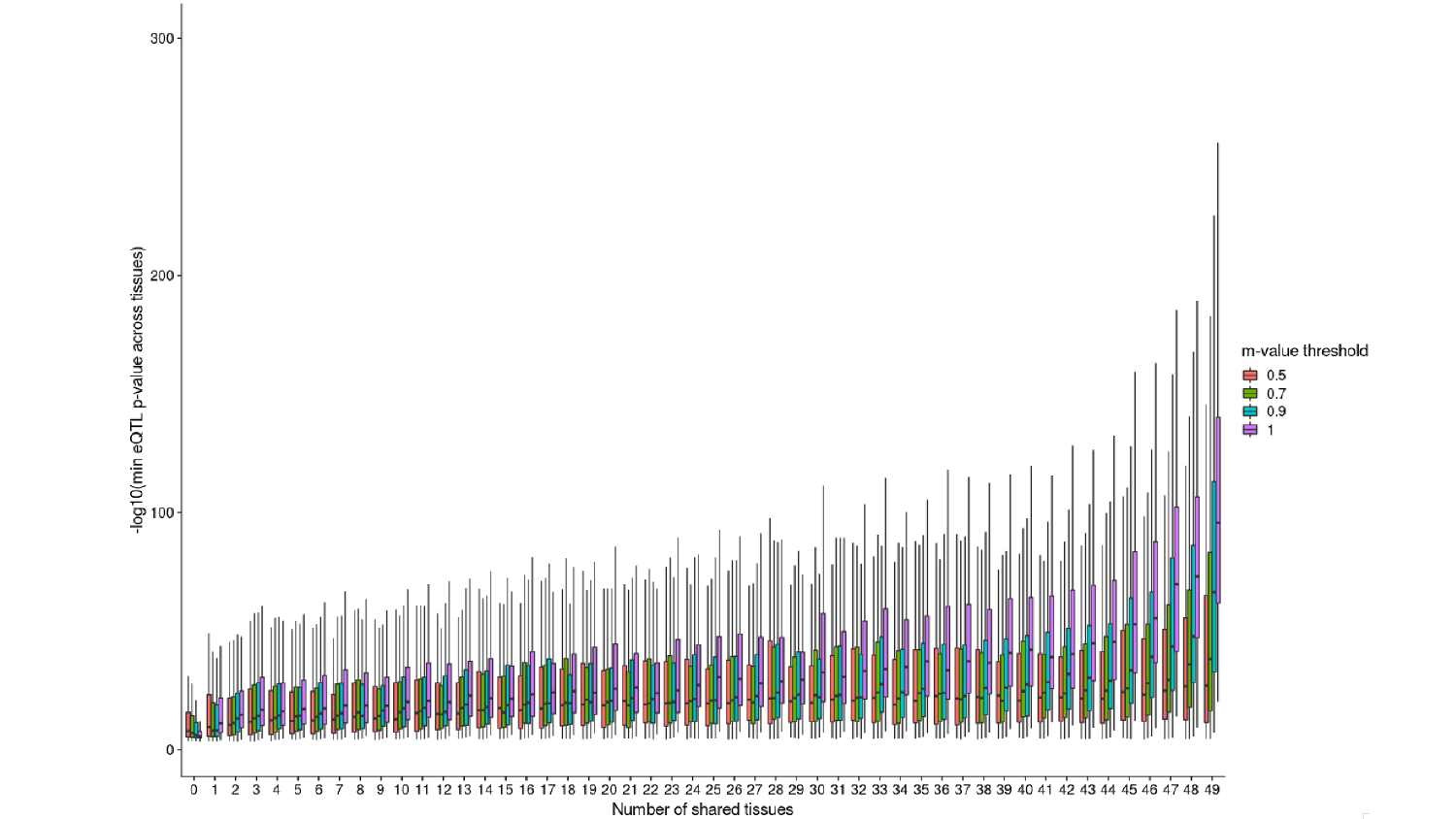

### Slide 8
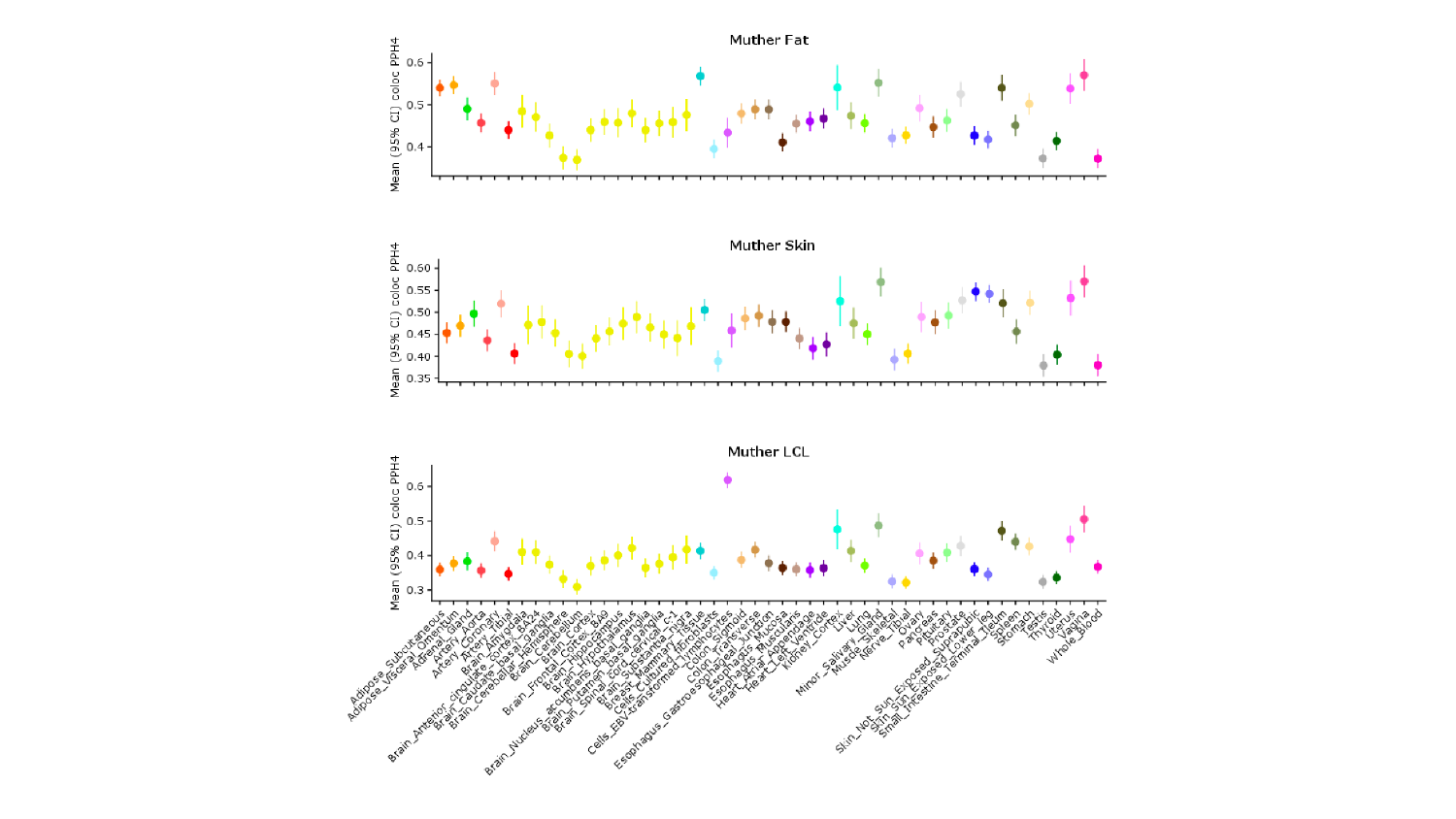

### Slide 9
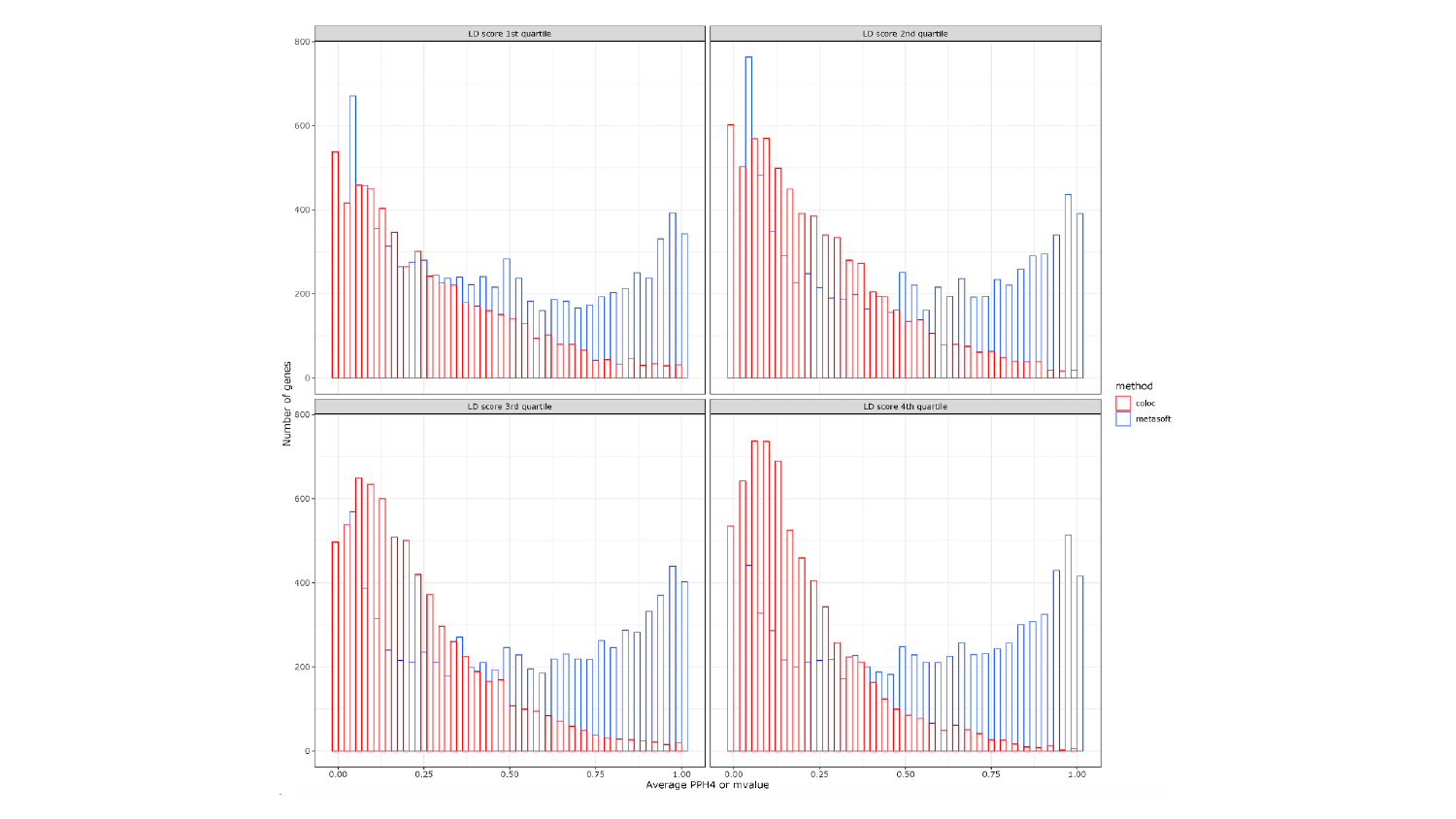

### Slide 10
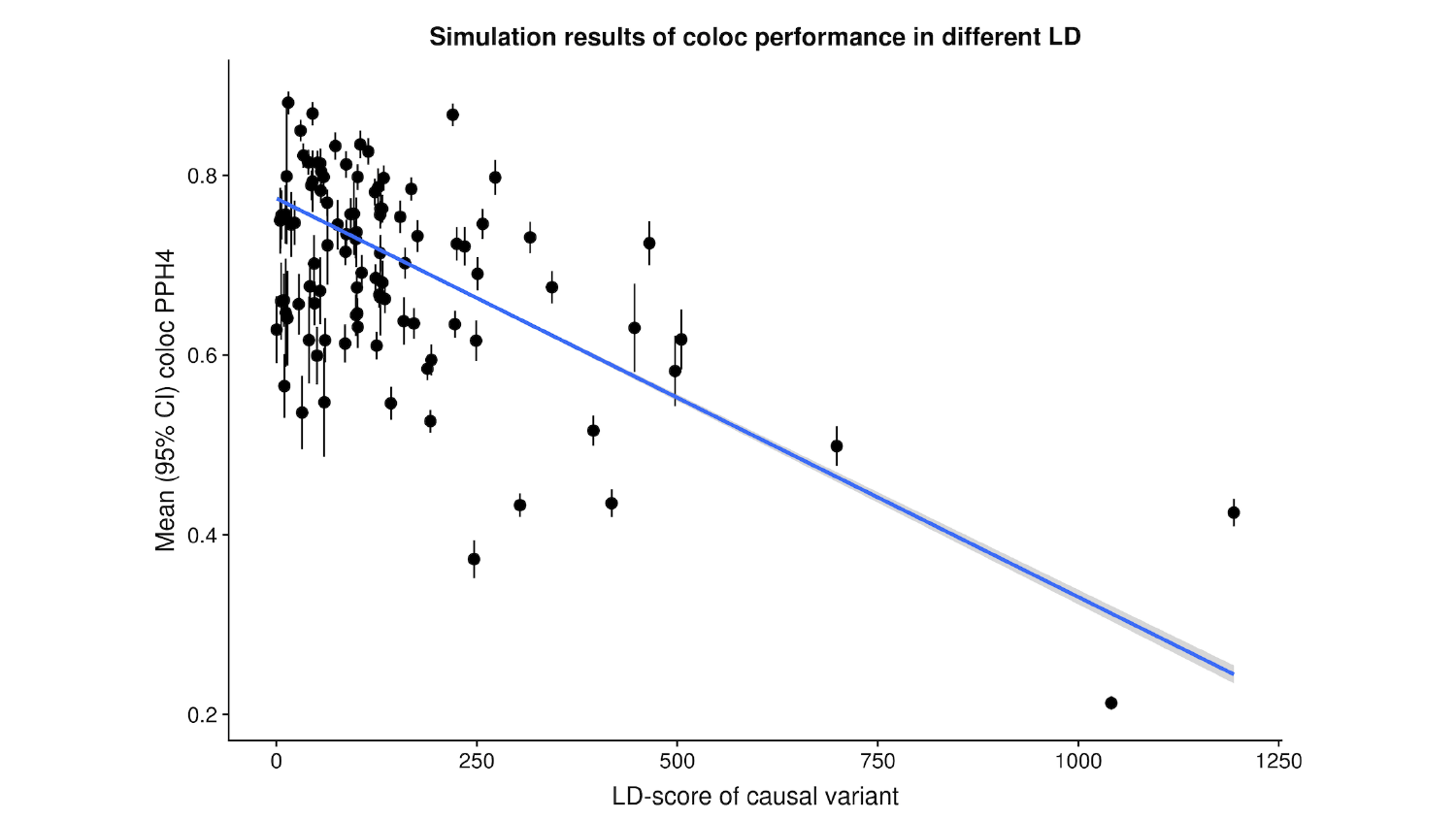

### Slide 11
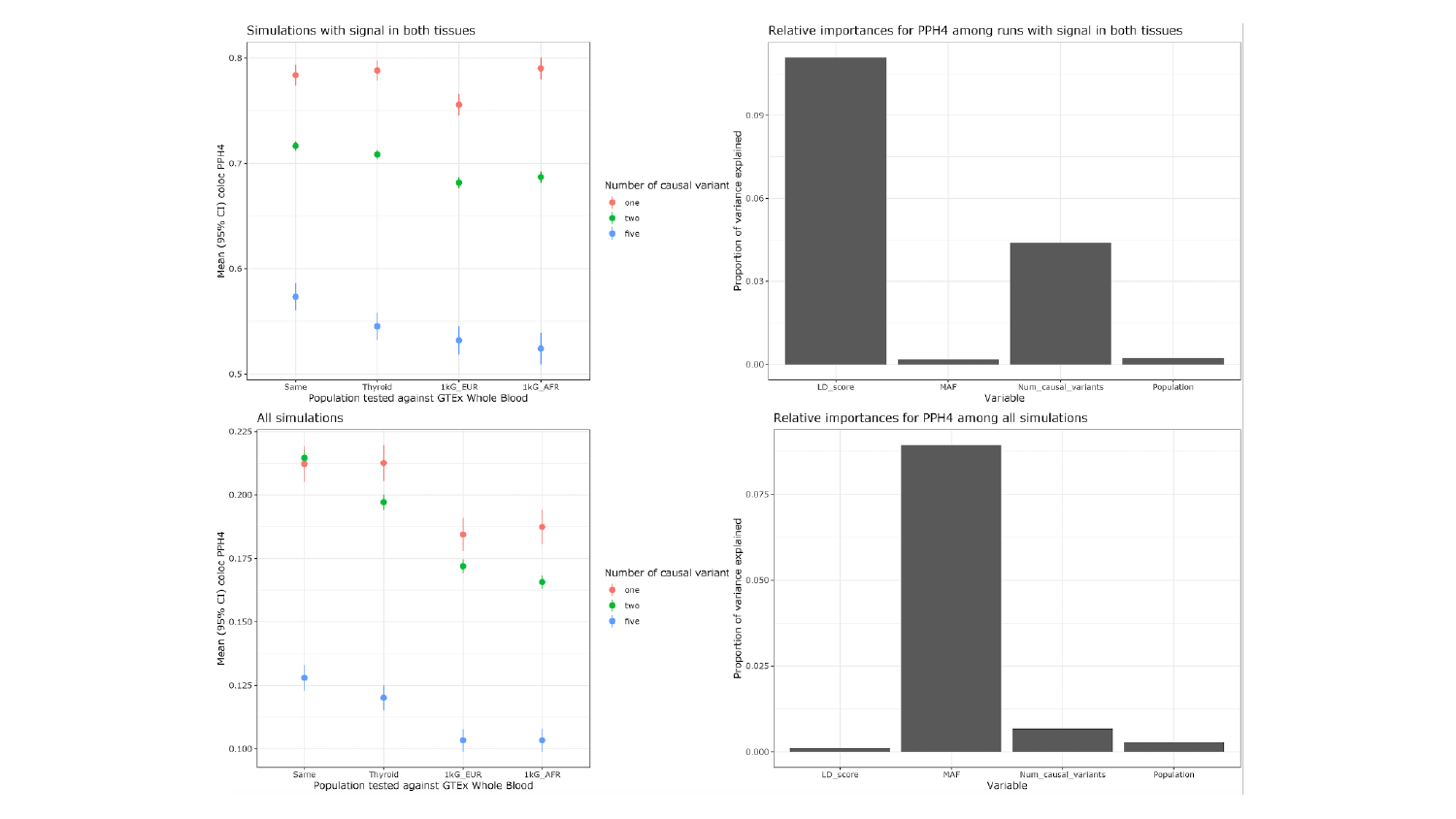

### Slide 12
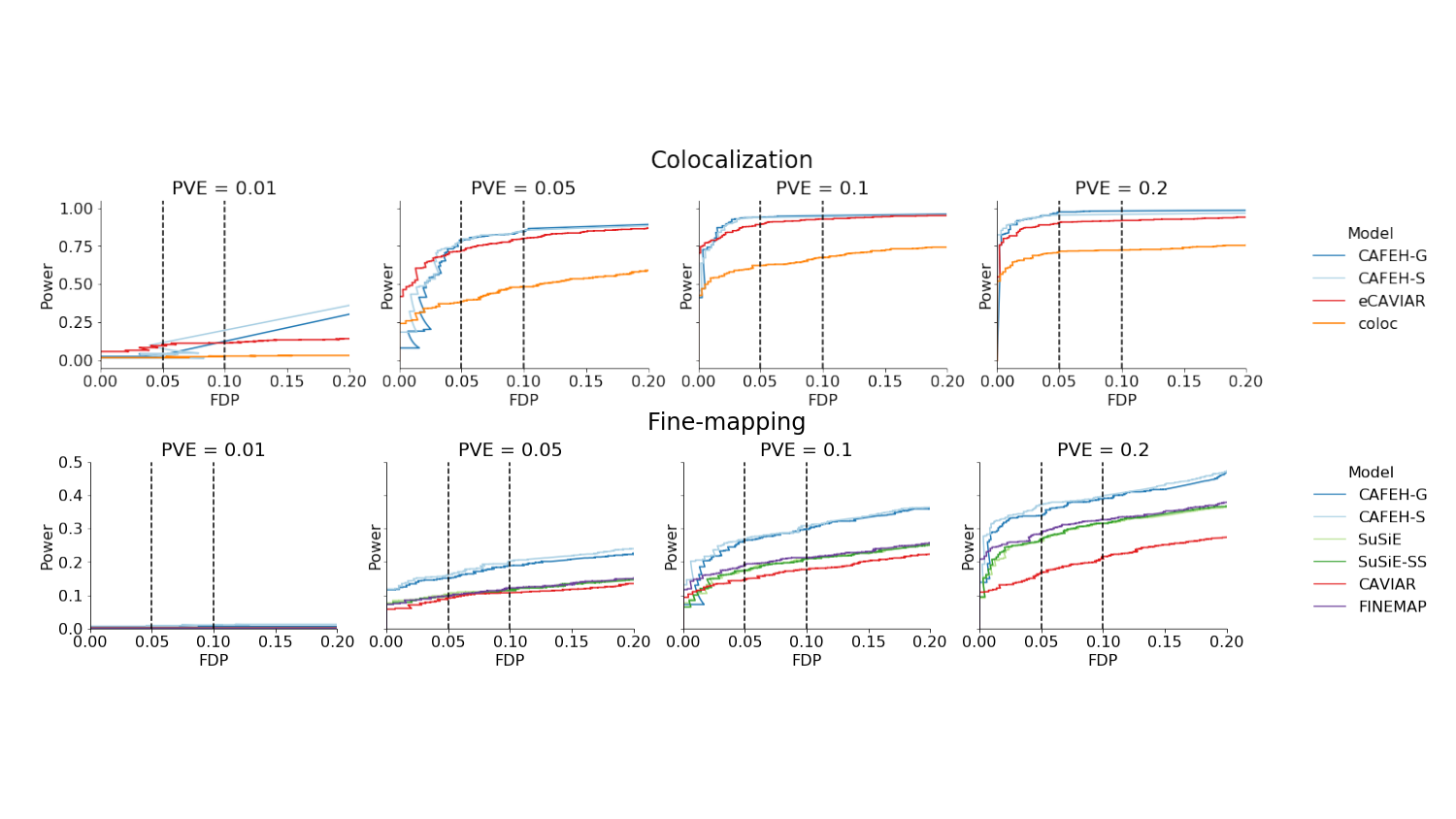

### Slide 13
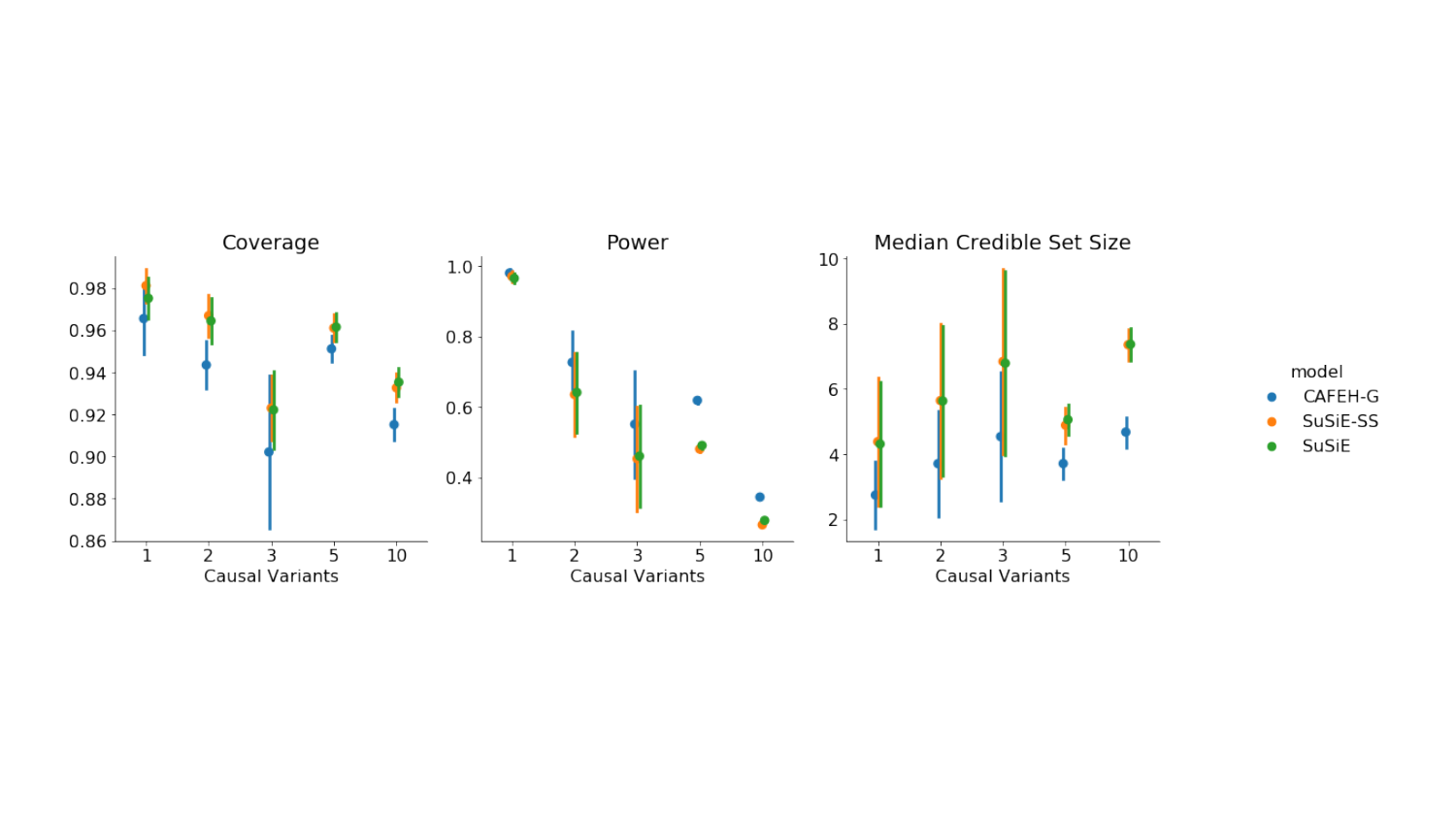

### Slide 14
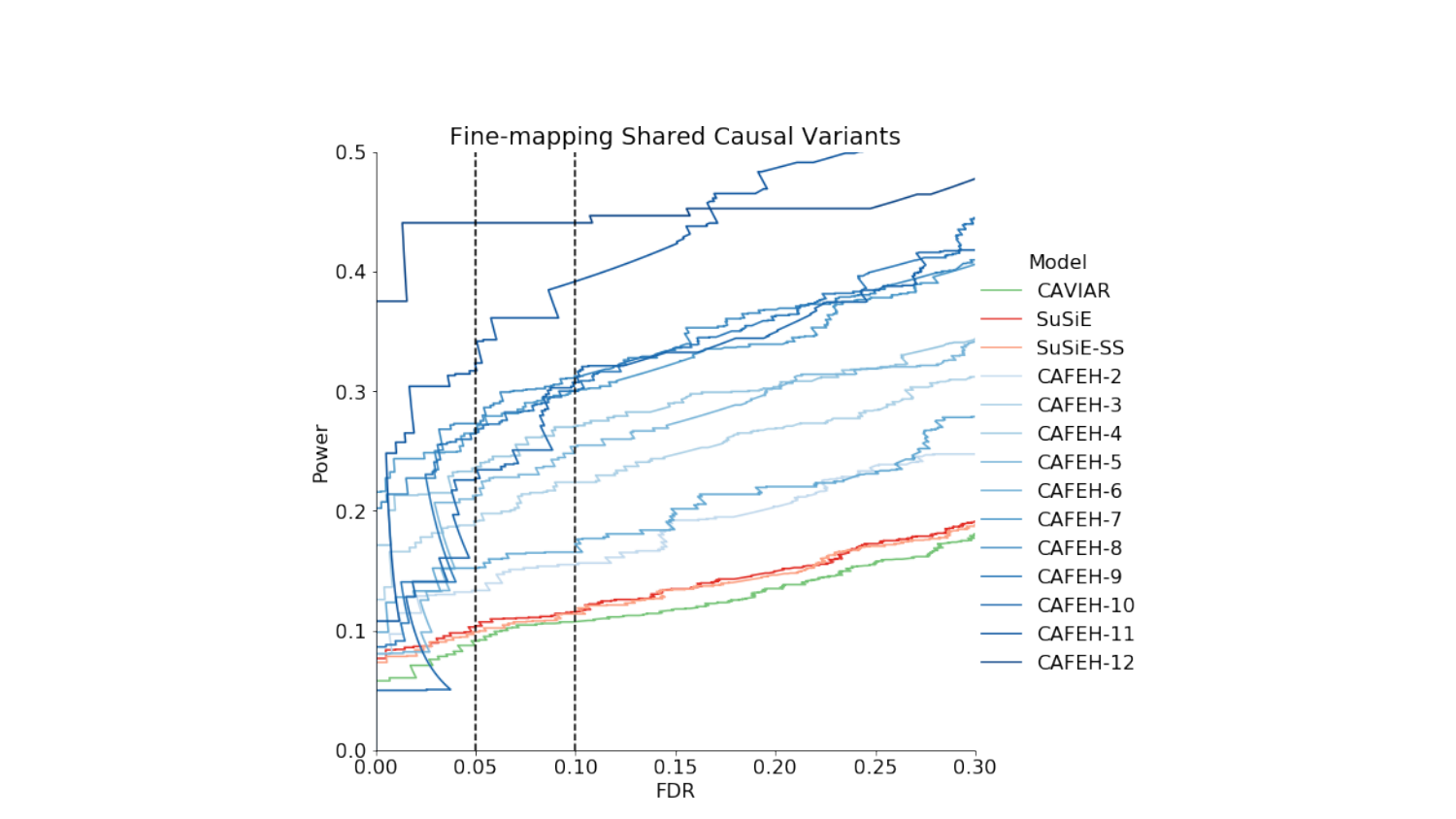

### Slide 15
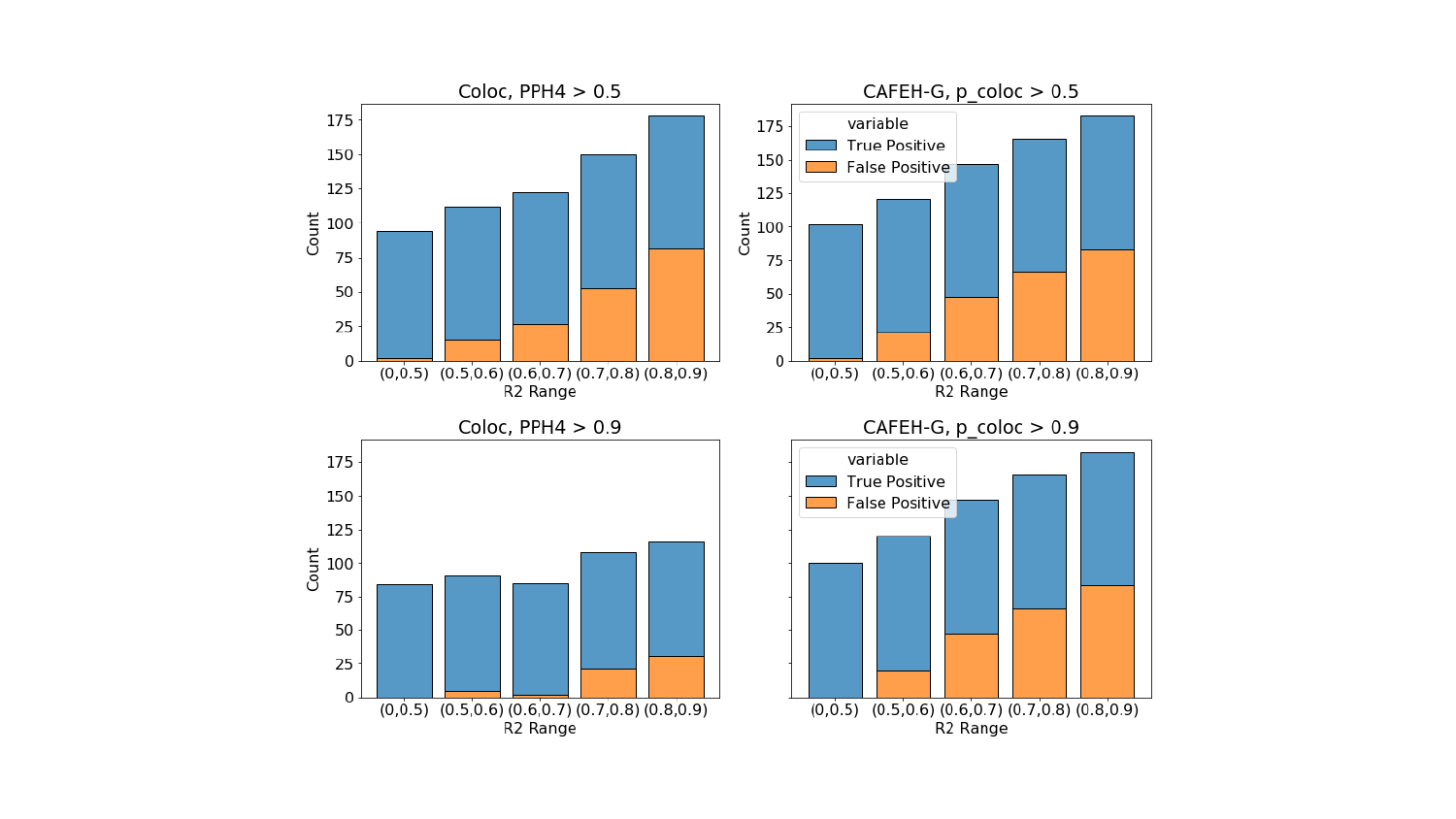

### Slide 16
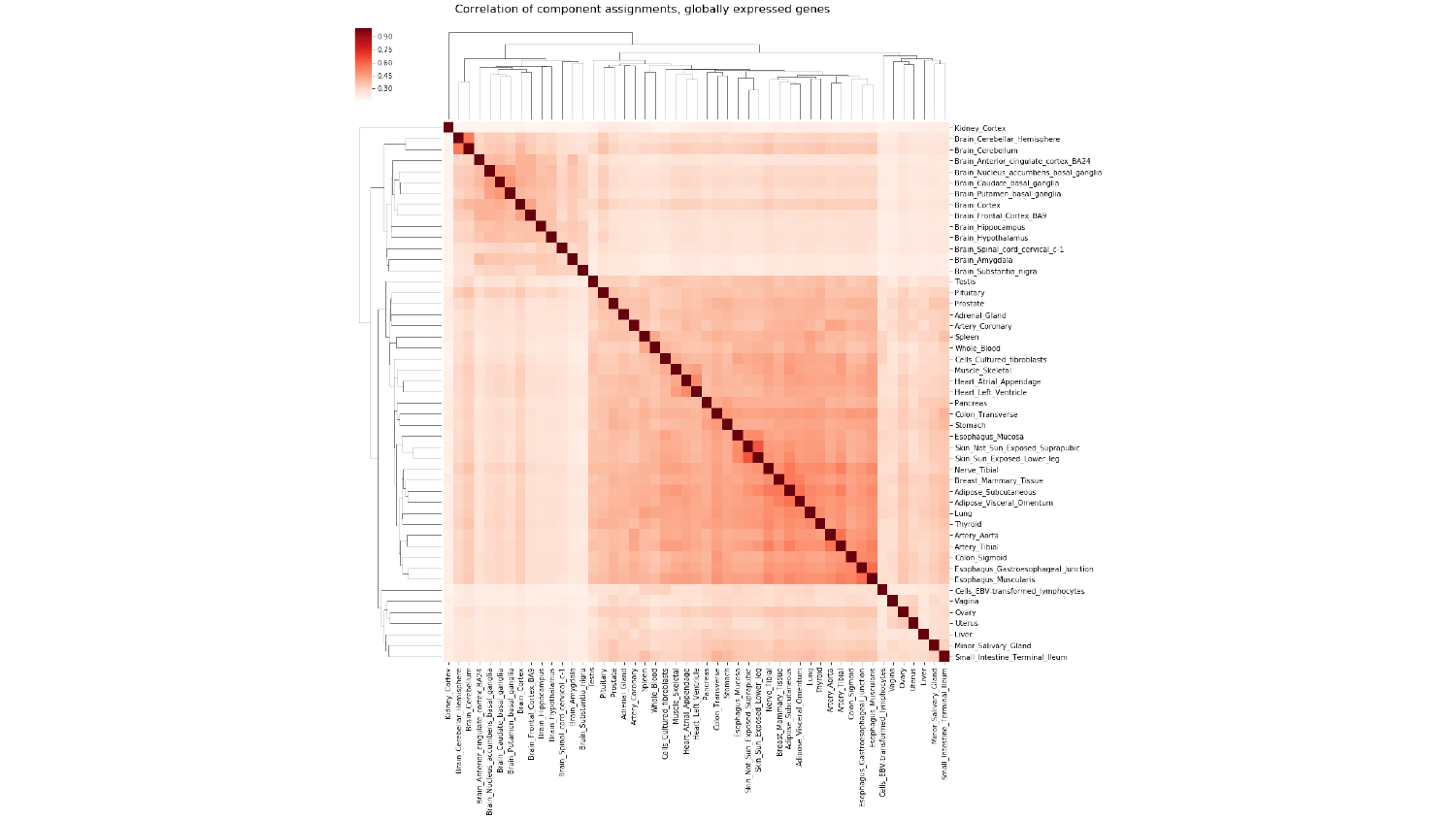

### Slide 17
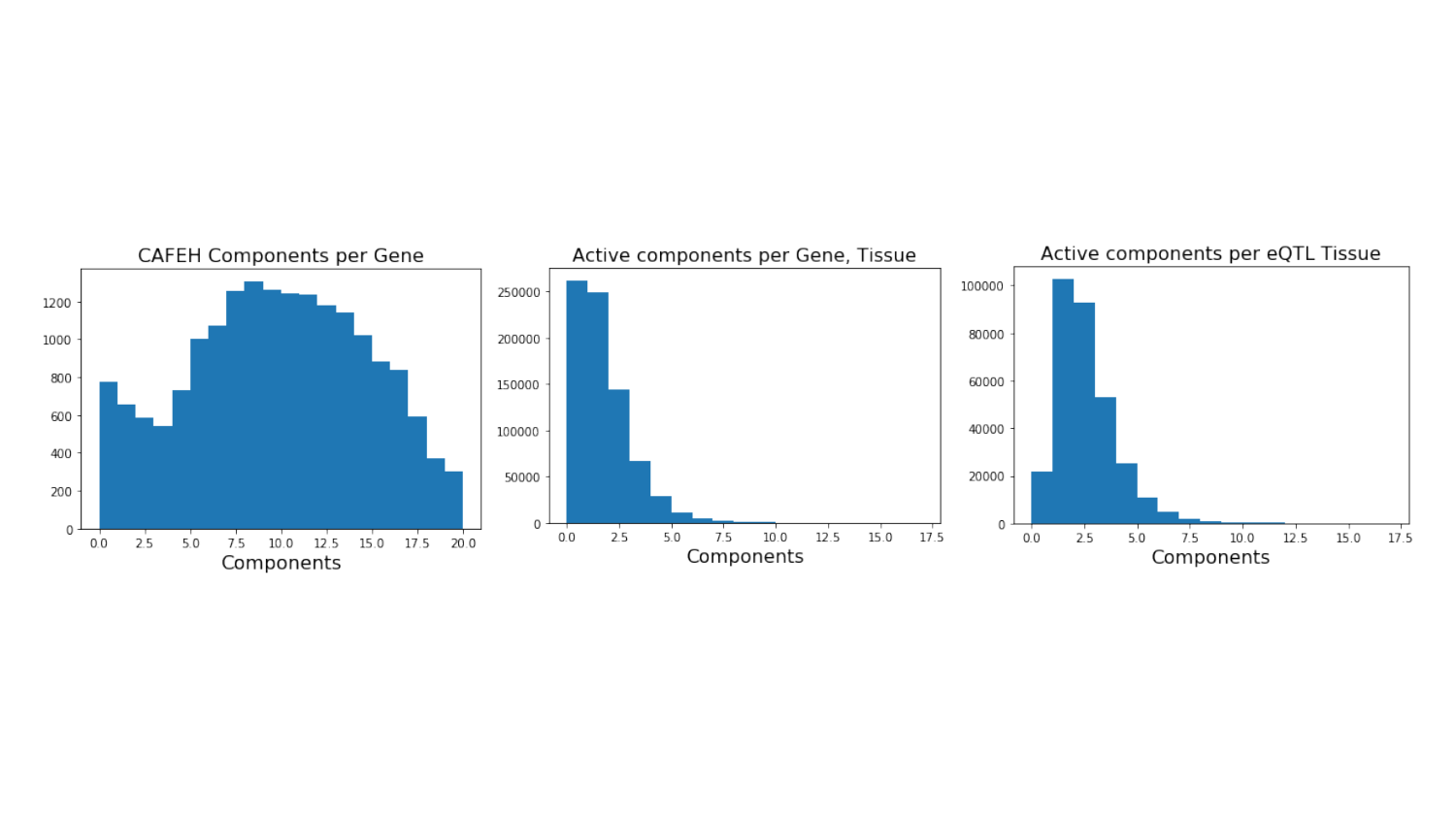

### Slide 18
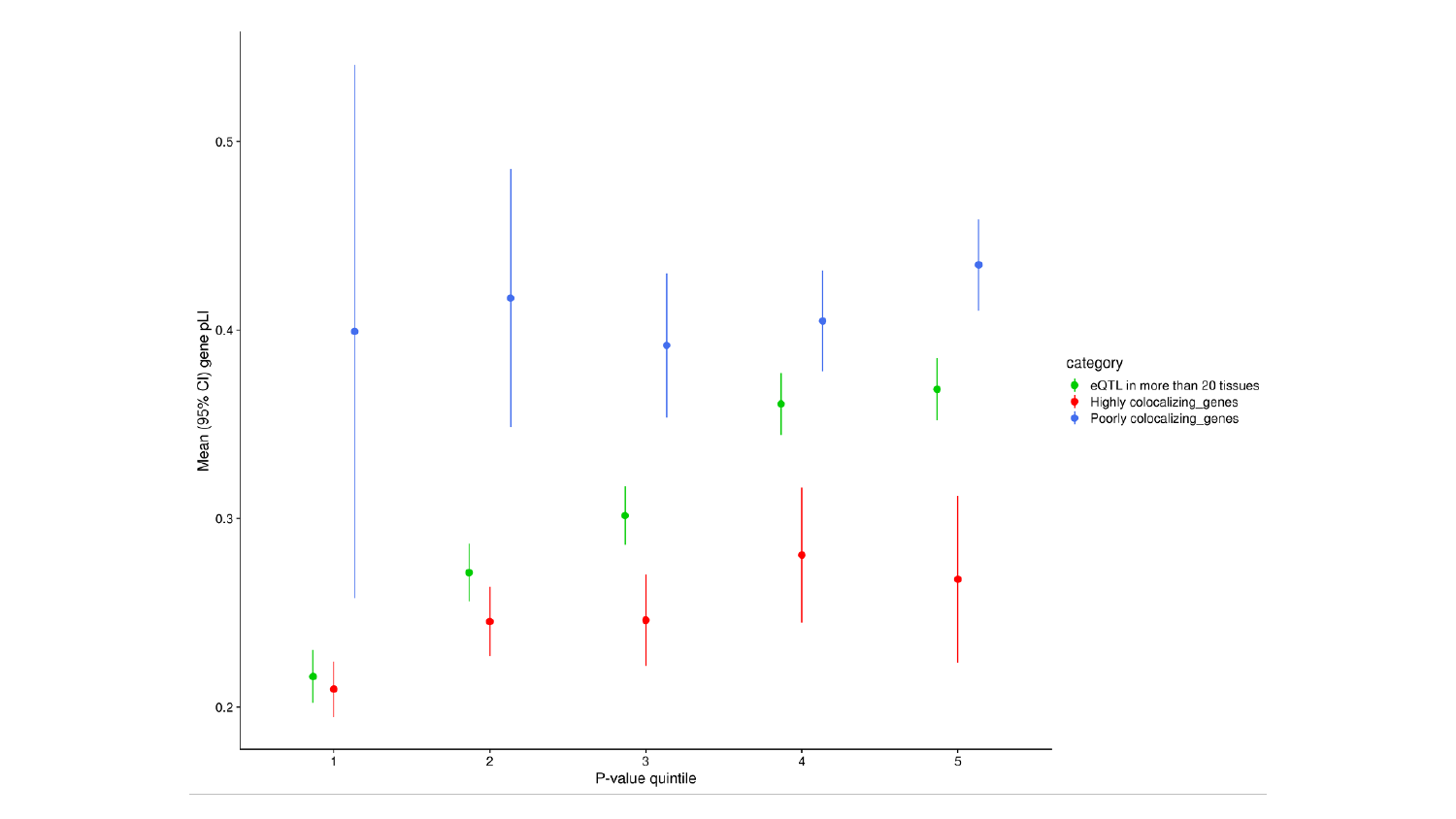

### Slide 19
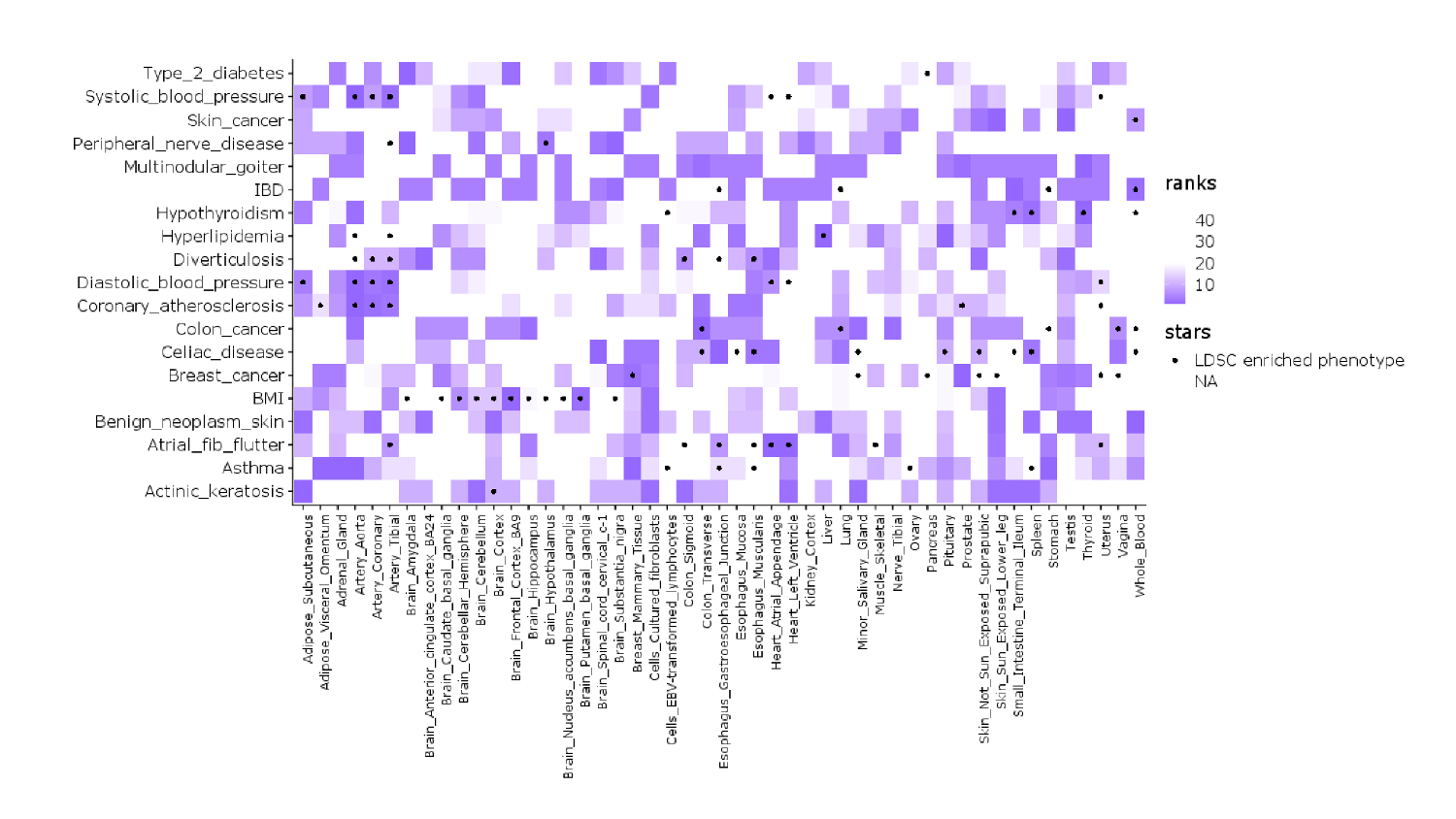

### Slide 20
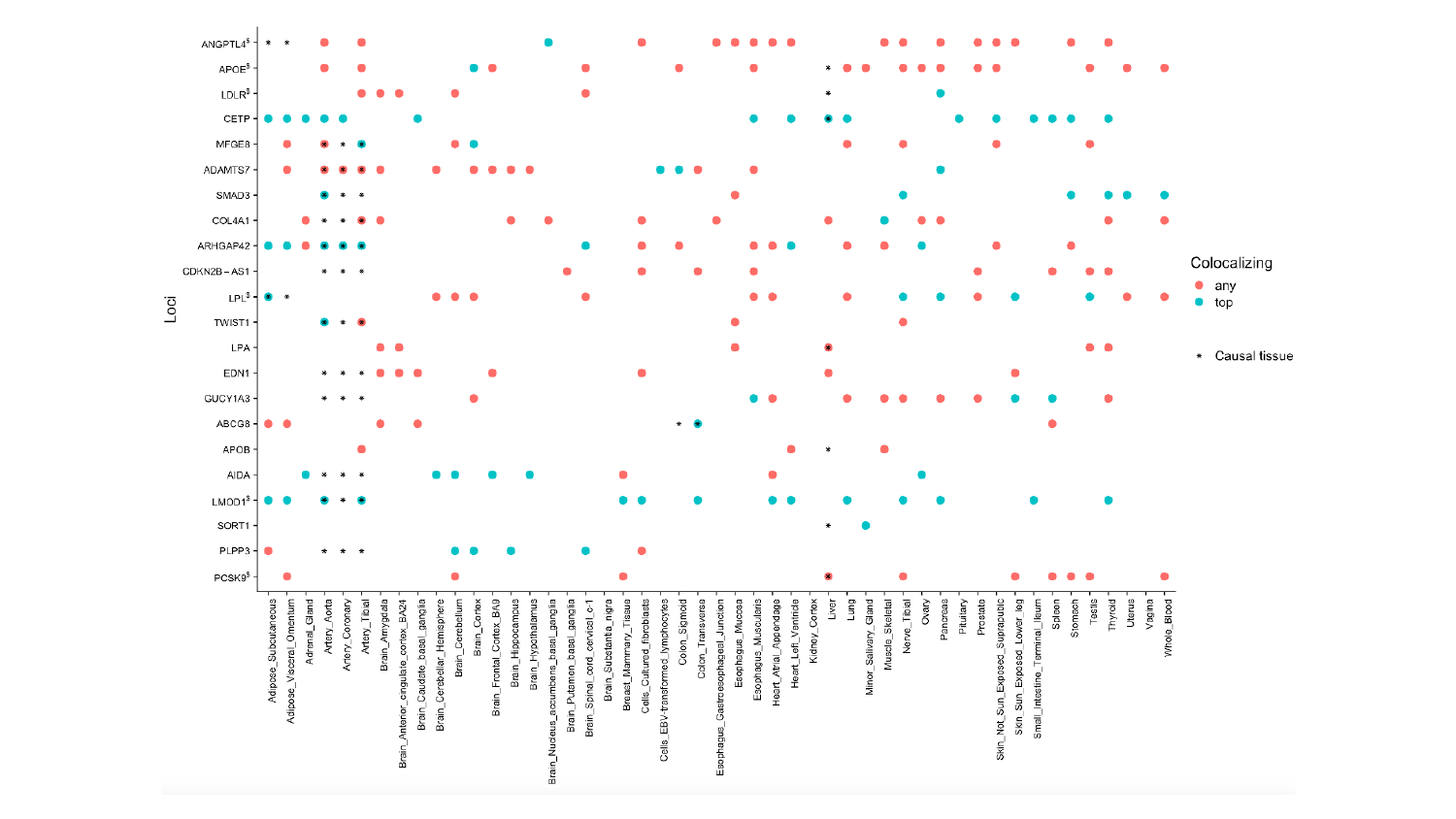
